# Mendelian Randomization Studies of Myopia: Choosing the right Summary Statistics

**DOI:** 10.1101/2025.06.04.25328975

**Authors:** Thu-Nga Nguyen, Louise Terry, Jeremy A. Guggenheim

## Abstract

**Purpose:** To examine if the choice of genome-wide association study (GWAS) summary statistics can yield invalid or misleading conclusions in Mendelian randomization (MR) studies of myopia.

**Methods:** The relationships between (1) years of full-time education and myopia, and (2) myopia and primary open-angle glaucoma (POAG), were used as exemplar testcases. MR analyses were performed with nine different sets of summary statistics for myopia: seven from sources widely used in published MR studies, plus two newly derived sets (a GWAS in either 66,773 unrelated participants or 93,036 participants that included relatives).

**Results:** Using the two newly derived sets of summary statistics from GWAS for myopia in unrelated and related samples, MR analyses demonstrated the expected positive causal relationship between education and myopia: odds ratio (OR) for myopia = 1.18, 95% confidence interval (CI) = 1.10 to 1.26 and OR = 1.16, 95% CI = 1.09 to 1.23 per additional year of education, respectively, and the expected positive relationship between myopia and POAG: OR = 1.11, 95% CI = 1.03 to 1.19 and OR = 1.12, 95% CI = 1.03 to 1.21, respectively. MR analyses performed using existing published GWAS summary statistics yielded highly inconsistent results, including MR estimates that suggested education protected against myopia and that myopia reduced the risk of POAG.

**Conclusions:** Care is required when designing MR analyses. Our findings imply that the results of some past MR studies of myopia were invalid.

## Introduction

Eye diseases have become increasingly prevalent, with detrimental effects on individuals, healthcare systems, and society.^1, 2^ For example, myopia now affects approximately one-third of the global population.^3^ Myopia increases the risk of sight-threatening eye disorders and results in an estimated productivity loss of around US$250 billion per year, globally.^4–9^ To address the burden of eye diseases, it is imperative to develop comprehensive preventive and management strategies by elucidating the risk factors and pathophysiological mechanisms underlying them.

Mendelian randomization (MR) is a statistical method for investigating the causal relationship between a modifiable environmental exposure and an outcome, relying solely on observational data.^10^ MR takes advantage of the random assortment of alleles during gametogenesis to define groups that, on average, differ in their level of the exposure-of-interest: Individuals who inherit “high-risk” alleles tend to experience a higher level of the exposure in comparison to those who inherit “low-risk” alleles. The random assortment of alleles during meiosis (Mendel’s second law) ensures that this genetically-conferred level of exposure is largely independent of socio- economic and other environmental or lifestyle risk factors (but this can only be guaranteed in the absence of population stratification, assortative mating, and genetic nurture^11^). If strict, so-called “instrumental variable”, assumptions are met, then MR is robust to the presence of confounding factors and reverse causation. Numerous vision-related studies have utilized MR to assess the causal relationship between variables, such as years of education and myopia risk,^12, 13^ myopia and primary open-angle glaucoma,^14, 15^ and many more.^16–24^

One of the fundamental steps in conducting a two-sample MR analysis is to select appropriate data sources. Summary statistics from published genome-wide association studies (GWAS) that include regression coefficients for single nucleotide polymorphism (SNP)-exposure or SNP-outcome relationships are readily available for download from public databases, repositories, or research consortia websites. These sets of GWAS summary statistics enable MR analyses to be conducted in a short time with minimal resource requirements, which has led to a rapid expansion of MR publications in the research literature.^10, 25^ Comprehensive guidelines on how to conduct or report MR studies have been published.^25, 26^ However, these guidelines have focused little attention on the importance of choosing appropriate data sources for an MR analysis. Given the availability of multiple data repositories, researchers typically face the question of which set of summary statistics for an outcome or exposure should they choose? The various GWAS analyses for a trait-of-interest may have used different participant cohorts, case definitions, or analysis methods. Each of these parameters will influence the estimated SNP-trait regression coefficients. This source of variability will feed through to influence the causal effect estimate of the MR analysis, potentially resulting in spurious findings that compromise the robustness and reliability of the research literature. The aim of the current work was to investigate if the choice of GWAS summary statistics can influence MR estimates. We focused on two exemplar testcases: (1) a two-sample MR analysis examining the role of educational attainment on myopia, and (2) a two-sample MR analysis examining the risk of primary open-angle glaucoma (POAG) in myopic vs. non-myopic individuals. Our findings advocate for a more scientifically rigorous approach in selecting summary statistics for MR and interpreting the results obtained from such studies.

## Methods

### Study cohorts

We used publicly available summary statistics of GWAS from four sources: the UK Biobank, the FinnGen study, the Social Science Genetic Association Consortium (SSGAC), and an international consortium of glaucoma genetics researchers^27^, as well as performing two new GWAS analyses for myopia in UK Biobank participants.

*UK Biobank*: Approximately 500,000 adults aged between 40 and 69 were recruited from 2006 to 2010.^28^ Participants visited one of 22 assessment centers across Great Britain for baseline and follow-up evaluations, during which their sociodemographic and clinical information was collected, including ophthalmic assessments completed by approximately 23% of participants. Genotype data were obtained using either the Biobank Axiom array (Affymetrix, High Wycombe, UK) or the BiLEVE Axiom array (Affymetrix), followed by imputation.

*FinnGen*: Approximately 500,000 individuals from Finland, averaging 53 years of age, consented to the use of their electronic health record (EHR) information and biological samples, as part of disease-based and population-based studies. The FinnGen cohort has been intentionally enriched with individuals suffering from various diseases. Phenotype data, which includes International Classification of Diseases (ICD)-10 codes, primarily derive from the Finnish national health registers, given Finland’s comprehensive population-wide registry coverage.

Genotype data were generated using the FinnGen ThermoFisher Axiom custom chip array (versions 1 and 2).^29^

*SSGAC*: A total of 293,723 adults of European ancestry from different cohorts with educational attainment information were included in a large GWAS meta-analysis conducted by Okbay et al.^30^ The primary outcome variable, EduYears, was imputed based on the participants’ years of schooling, which was standardized according to the 1997 International Standard Classification of Education.

*Gharahkhani et al.*^27^ *glaucoma genetics consortium*. A GWAS meta-analysis of POAG was conducted in 216,257 adults of European ancestry. POAG diagnosis was based on ICD9/ICD10 criteria. Ethical approval for the study was obtained from each of the participating institutions. All participants provided informed consent.

### GWAS summary statistics for years of education

Summary statistics for the SSGAC GWAS for EduYears (n=293,723) were reported by Okbay et al.^30^ The SNP-EduYears regression coefficients were reported in units of standard deviation in the original paper; for the current analyses, these were converted to units of years of full-time education using a conversion factor of 1 standard deviation = 3.6 years in school.^30^

### GWAS summary statistics for primary open-angle glaucoma

Summary statistics for a GWAS meta-analysis of POAG across 21 independent samples of European ancestry (n = 16,677 POAG cases and n = 199,580 controls) were reported by Gharahkhani et al.^27^ UK Biobank participants (n = 1,448 cases and n = 22,107 controls) and FinnGen participants (n = 1,824 cases and n = 93,036 controls) were included in the Gharahkhani et al. GWAS meta-analysis of POAG. However, this small degree of sample overlap with UK Biobank and FinnGen was not expected to appreciably bias 2-sample MR analyses.^31^

### GWAS summary statistics for myopia

Publicly available GWAS summary statistics for myopia in European-ancestry individuals were identified by searching the following public databases: GeneATLAS (Roslin Institute and Medical Research Council Human Genetics Unit, University of Edinburgh), GWASATLAS (VU University of Amsterdam), GWAS Catalog (National Human Genome Research Institute, European Molecular Biology Laboratory – European Bioinformatics Institute), IEU OpenGWAS project (UK Medical Research Council Integrative Epidemiology Unit [IEU], University of Bristol), and FinnGen. To facilitate the search, the terms “myopia,” “nearsightedness,” “shortsightedness,” and “refractive error” were employed in the database queries. The results are presented in Table 1. The majority of published MR studies used GWAS summary statistics for analyses conducted in the UK Biobank cohort, making use of participants’ self-reported “Reason for glasses/contact lens: For short-sightedness (called ‘myopia’)” (UK Biobank data field 6147; response option 1). The remainder of the published studies used GWAS summary statistics from analyses conducted in FinnGen, which used the ICD code H52.1 or Phecode 367.1 to define myopia. Ultimately, seven existing sets of GWAS summary statistics were included in a series of MR analyses. Details of each dataset are provided in Table 1 and Supplementary Note S1.

**Table 1.** Publicly available and newly derived GWAS summary statistics for myopia.

| Database | ID/Authors | Study cohort | Myopia definition | Cases (N) | Controls (N) | Myopia prevalence in GWAS sample | Published MR study using the summary statistics |
| --- | --- | --- | --- | --- | --- | --- | --- |
| GWAS Catalog | GCST90044326<br>Jiang et al.<sup>39</sup> | UK Biobank | Reason for glasses/contact lenses: For short-sightedness (UKB data field 6147) | 36,623 | 419,031 | 8.0% | - |
| GWAS Catalog | GCST90435990<br>Zhou et al.<sup>33</sup> | UK Biobank | Myopia (PheCode 367.1) | 1,257 | 406,530 | 0.3% | - |
| IEU OpenGWAS | ukb-a-419<br>(Neale Lab) | UK Biobank | Reason for glasses/contact lenses: For short-sightedness (UKB data field 6147) | 26,943 | 308,757 | 8.0% | Xu et al.<sup>54</sup> |
| IEU OpenGWAS | ukb-b-6353 | UK Biobank | Reason for glasses/contact lenses: For short-sightedness (UKB data field 6147) | 37,362 | 423,174 | 8.1% | Deng et al.<sup>55</sup><br>Jiang et al.<sup>56</sup><br>Li et al.<sup>57</sup><br>Liang et al.<sup>58</sup><br>Lv et al.<sup>59</sup><br>Mo et al.<sup>60</sup><br>Wei et al.<sup>61</sup><br>Xu et al.<sup>54</sup><br>Xu et al.<sup>62</sup><br>Zhang et al.<sup>63</sup><br>Zhu et al.<sup>64</sup> |
| FinnGen (R5) | finn-b-H7_MYOPIA | FinnGen | ICD Code (H52.1) | 1,640 | 210,931 | 0.8% | Li et al.<sup>65</sup><br>Wei et al.<sup>66</sup><br>Wei et al.<sup>61</sup><br>Wei et al.<sup>67</sup><br>Xu et al.<sup>54</sup><br>Zhu et al.<sup>64</sup> |
| FinnGen (R9) | H7_MYOPIA | FinnGen | ICD Code (H52.1) | 3,534 | 361,237 | 1.0% | Deng et al.<sup>55</sup> |
| FinnGen (R10) | finn-b-H7_MYOPIA | FinnGen | ICD Code (H52.1) | 4106 | 394,028 | 1.0% | Hui et al.<sup>68</sup><br>Huang et al.<sup>69</sup> |
| Newly-derived | Current study | UK Biobank | SER in at least 1 eye ≤ -0.5 diopter (including related individuals) | 35,531 | 57,505 | 38.2% | - |
| Newly-derived | Current study | UK Biobank | SER in at least 1 eye ≤ -0.5 diopter (only unrelated individuals) | 25,804 | 40,969 | 38.6% | - |

Abbreviations: GWAS= Genome wide association study; ICD=International Classification of Diseases; SER=Spherical equivalent refractive error

### Newly performed GWAS for myopia in UK Biobank

Given the limitations of the currently available GWAS summary statistics for myopia (see below), we performed two new GWAS analyses for myopia in UK Biobank participants of European ancestry who had non-cycloplegic autorefraction measurement data and no other eye disorders. Details of the GWAS analyses can be found in Supplementary Note S2. Briefly, spherical equivalent refractive error (SER) was calculated as the autorefraction sphere power plus half the cylinder power. Participants were classified as myopia cases if the SER ≤ -0.50 diopters (D) in at least one eye. Controls were classified as individuals with SER > -0.50 D in both eyes. A GWAS for myopia in unrelated individuals (n = 25,804 cases; n = 40,969 controls) was conducted using PLINK2,^32^ while a GWAS for myopia that included related individuals (n = 35,531 cases; n = 57,505 controls) was conducted using SAIGE/GATE.^33^ Summary statistics for the newly performed GWAS analyses are available at: doi: 10.17035/cardiff.29211071. Ethical approval for the UK Biobank study was obtained from the Northwest Multicentre Research Ethics Committee (Reference: 11/NW/0382). Participants provided informed consent and were free to withdraw from the study at any time. The research adhered to the tenets of the Declaration of Helsinki.

### Selection of instrumental variables for years of education

We used summary statistics from the Okbay et al.^30^ GWAS for EduYears discovery analysis to avoid overlap with UK Biobank. GWAS variants independently associated with EduYears at P < 5 x 10^-8^ were selected; subsequently, variants not included in the Haplotype Reference Consortium (HRC) were excluded, as were SNPs not present in any of the myopia GWAS datasets, to ensure consistency in the genetic instrumental variables utilized across all of the MR analyses. Ultimately, 62 genetic variants remained suitable for use as instrumental variables (Supplementary Table S1).

### Selection of instrumental variables for myopia

We used a consistent approach to select IVs for myopia from each of the nine sets of GWAS summary statistics for myopia listed in Table 1. We first excluded variants not available in the summary statistics for POAG^27^ or not available in the clumping reference panel (N = 10,000 UK Biobank participants of European ancestry). Variants were clumped using PLINK v1.9 using a p-value threshold of *P* < 5.0e-08, a distance metric of ±1000 kb and a linkage disequilibrium threshold of *r*^2^ < 0.05.^32^

### Mendelian randomization analysis

Full details of the MR analyses can be found in Supplementary Note S3 and code to reproduce the analyses is provided in Supplementary Note S4. The inverse-variance weighted (IVW) MR method was chosen as the primary analysis method.^34^ The following sensitivity analyses were performed to evaluate the robustness of the IVW-MR analysis against the assumption of no horizontal pleiotropy: MR-EGGER^35^, weighted median MR^36^, mode-based MR^37^, and MR PRESSO^38^.

## Results

A search of online GWAS repositories yielded 16 sets of myopia summary statistics publicly available for download. Twelve of the 16 sets of summary statistics were from the FinnGen study. We included the myopia summary statistics from FinnGen release 5, 9 and 10 in the current work, since these 3 sets have been utilized in published MR studies of myopia (Table 1). Two of the 16 sets of myopia summary statistics were from the IEU OpenGWAS database; we included these as they have been very widely used in published MR studies of myopia (Table 1). Finally, 2 of the 16 sets of myopia summary statistics were from the GWAS Catalog. Although these have not been used in published MR studies to our knowledge, we included them in the current analyses because their convenient availability would make them potentially suitable for an MR study of myopia.

To provide gold standard MR causal effect estimates to compare against those obtained using the publicly available myopia summary statistics, we performed two new GWAS analyses for myopia in samples of UK Biobank participants whose SER had been measured by autorefraction. These new sets of summary statistics have been made openly accessible.

### Mendelian randomization analysis of the relationship between years of education and myopia

An IVW-MR analysis using summary statistics from our newly-performed GWAS for myopia in unrelated participants (25,804 cases and 40,969 controls) yielded an estimate for the causal effect of education on myopia of OR = 1.18 per year of education (95% CI 1.10 to 1.26; *P* = 3.1e-06). An IVW-MR analysis using summary statistics from our newly-performed GWAS for myopia that included related individuals (35,531 cases and 57,505 controls) yielded a similar causal effect estimate of OR = 1.16 per year of education (95% CI 1.09 to 1.23; *P* = 2.2e-06).

The results of the IVW-MR analyses using the seven sets of publicly-available GWAS summary statistics are presented in Table 2, Figure 1 and Supplementary Figure S1. Complete results using the full range of MR methods are presented in Supplementary Table S2. These MR analyses yielded varying causal effect estimates for the relationship between education and myopia. As well as variability in effect size, the level of statistical significance of the causal effect varied widely, too. For instance, the MR analysis using GWAS summary statistics ukb-a-419 from the Neale lab repository suggested a small but highly significant *negative* causal relationship between years spent in full-time education and myopia status (OR = 0.99 per year of education, 95% CI = 0.988 to 0.996, *P* = 1.7e-05), implying that additional education was protective against myopia. GWAS summary statistics ukb-b-6353 from the IEU OpenGWAS project suggested evidence of a very small, highly significant positive causal effect of EduYears on myopia (OR = 1.01 per year of education, 95% CI = 1.006 to 1.014, *P* = 1.7e-07). The other set of GWAS summary statistics provided evidence for a positive causal effects ranging from OR = 1.07 to OR = 1.24.

**Figure 1:**
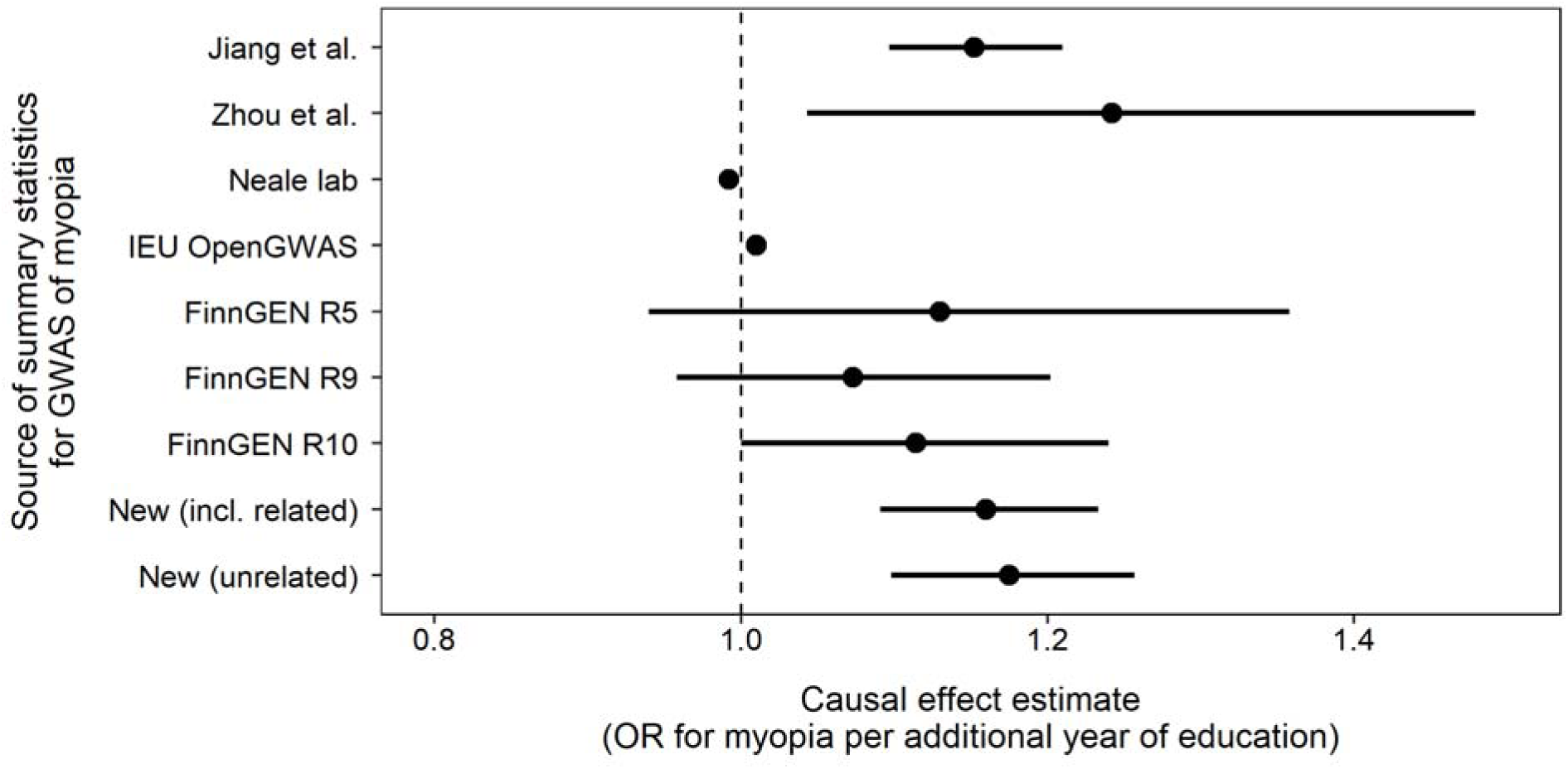
**Causal effect estimates from Mendelian Randomization analyses of education as a risk factor for myopia obtained using different sets of GWAS summary statistics for myopia**. Each data point represents an analysis using a different set of GWAS summary statistics for myopia. Error bars indicate 95% confidence intervals.

**Table 2.** Causal effect of years of education on the risk of myopia using different sets of summary statistics. Results were obtained using the IVW-MR method.

| **GWAS for myopia summary statistics** | **OR** | **95% CI** | **Effect\*** | **SE** | **P-value** |
| --- | --- | --- | --- | --- | --- |
| GCST90044326 (Jiang et al.<sup>39</sup>) | 1.152 | 1.097, 1.210 | 0.141 | 0.025 | 1.63e-08 |
| GCST90435990 (Zhou et al.<sup>33</sup>) | 1.242 | 1.043, 1.479 | 0.217 | 0.089 | 1.49e-02 |
| ukb-a-419 (Neale lab) | 0.992 | 0.988, 0.996 | -0.008 | 0.002 | 1.70e-05 |
| ukb-b-6353 (IEU OpenGWAS) | 1.010 | 1.006, 1.014 | 0.010 | 0.002 | 1.74e-07 |
| FinnGen (R5) | 1.130 | 0.940, 1.358 | 0.122 | 0.094 | 1.94e-01 |
| FinnGen (R9) | 1.073 | 0.958, 1.202 | 0.070 | 0.058 | 2.24e-01 |
| FinnGen (R10) | 1.114 | 1.000, 1.240 | 0.108 | 0.055 | 4.92e-02 |
| Newly derived (including related individuals) | 1.160 | 1.091, 1.233 | 0.148 | 0.031 | 2.15e-06 |
| Newly derived (unrelated individuals only) | 1.175 | 1.098, 1.257 | 0.161 | 0.035 | 3.11e-06 |

\*Estimated effect in units of log odds ratio for myopia per additional year spent in education.
Abbreviations: OR=Odds ratio; CI=Confidence interval; SE=Standard error; GWAS= Genome wide association study;

### Mendelian randomization analysis of the relationship between myopia and POAG

An IVW-MR analysis using summary statistics from the newly-performed GWAS for myopia in unrelated participants (25,804 myopia cases and 40,969 controls) yielded an estimate for the risk of POAG in myopic vs. non-myopic individuals of OR = 1.11 (95% CI = 1.03 to 1.19, P = 6.5e- 03). An IVW-MR analysis using summary statistics from the newly-performed GWAS for myopia that included relatives (35,531 myopia cases and 57,505 controls) yielded an estimate for the risk of POAG in myopic vs. non-myopic individuals of OR = 1.12 (95% CI = 1.03 to 1.21, *P* = 5.7e-03).

The results of the IVW-MR analyses using the publicly-available GWAS summary statistics are presented in Table 3, Figure 2 and Supplementary Figure S2. Complete results using the full range of MR methods are presented in Supplementary Table S3. Once again, MR analyses employing different sets of GWAS summary statistics for myopia yielded highly varied results. Strikingly, the MR analysis using myopia summary statistics ukb-a-419 from the Neale lab repository suggested myopia had a highly protective effect against POAG (OR = 0.07, 95% CI = 0.02 to 0.24, *P* = 4.1e-05). Equally striking was the estimated causal effect of myopia on POAG obtained with summary statistics ukb-b-6353 from the IEU OpenGWAS database, which suggested myopia significantly increased the risk of POAG several fold (OR = 8.98, 95% CI = 2.19 to 36.83, *P* = 2.3e-03). MR analyses using myopia summary statistics from Zhou et al.^33^ or FinnGen R9 and R10 suggested there was no evidence of a causal effect of myopia on POAG (all *P* > 0.05). Only the myopia summary statistics of Jiang et al.^39^ yielded results similar to those from the newly derived summary statistics: OR = 1.14, 95% CI = 1.04 to 1.25, *P* = 5.6e-03. The number of IVs for myopia in these MR analyses also varied widely (Table 3). Indeed, for the FinnGen R5 myopia summary statistics, no GWAS variant met our p-value threshold of *P* < 5.0e-08, hence there were no IVs available for an MR analysis using the FinnGen R5 summary statistics.

**Figure 2:**
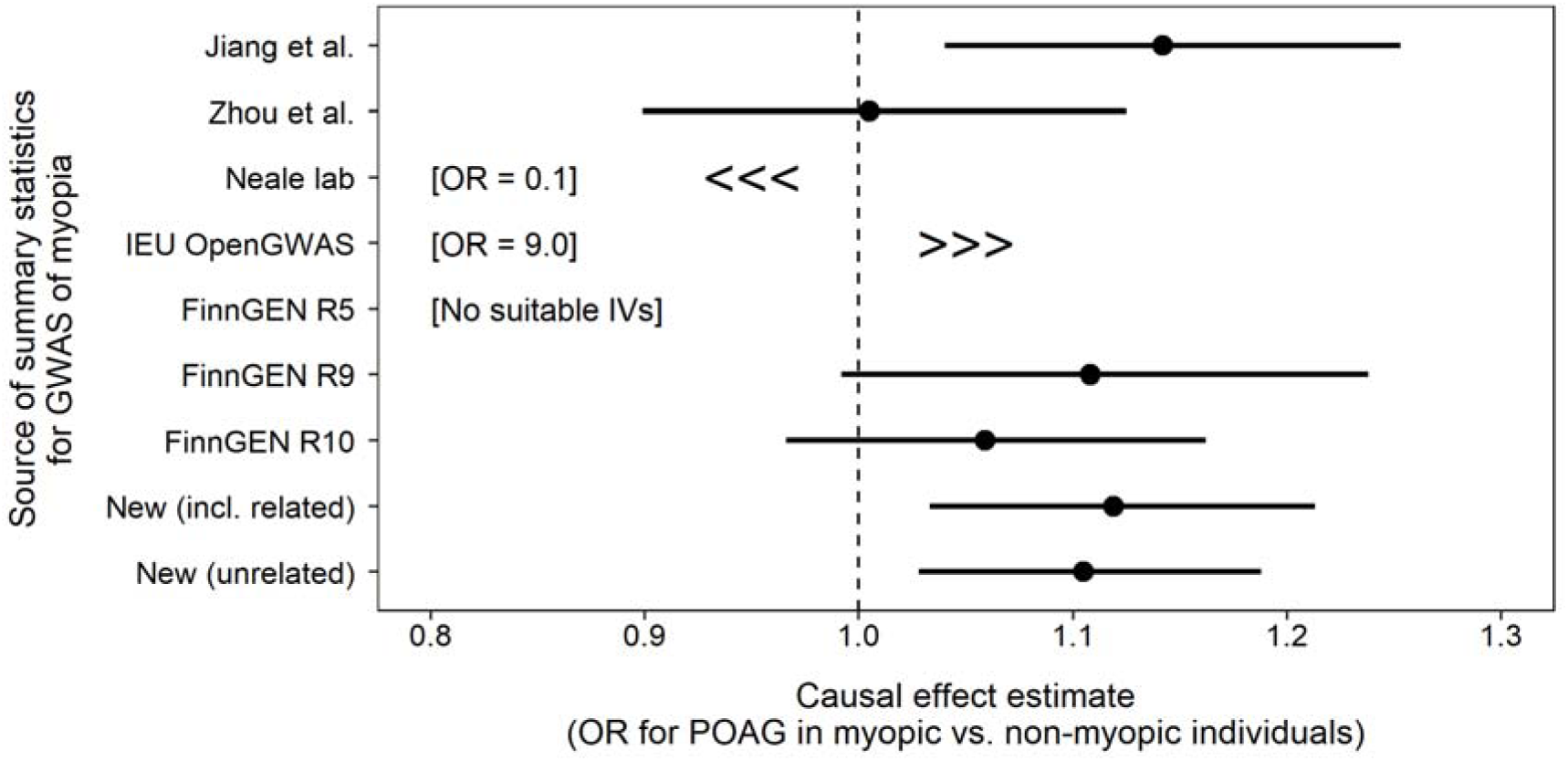
Causal effect estimates from Mendelian Randomization analyses of myopia as a risk factor for primary open-angle glaucoma obtained using different sets of GWAS summary statistics for myopia. Each data point represents an analysis using a different set of GWAS summary statistics for myopia. Error bars indicate 95% confidence intervals.

**Table 3.** Causal effect of myopia on the risk of primary open-angle glaucoma using different sets of summary statistics. Results were obtained using the IVW-MR method.

| **GWAS for myopia summary statistics** | **IVs (N)** | **OR** | **95% CI** | **Effect\*** | **SE** | **P-value** |
| --- | --- | --- | --- | --- | --- | --- |
| GCST90044326 (Jiang et al.<sup>39</sup>) | 32 | 1.142 | 1.040, 1.253 | 0.132 | 0.048 | 5.56E-03 |
| GCST90435990 (Zhou et al.<sup>33</sup>) | 1 | 1.005 | 0.899, 1.125 | 0.005 | 0.057 | 9.26E-01 |
| ukb-a-419 (Neale lab) | 17 | 0.066 | 0.018, 0.242 | -2.718 | 0.663 | 4.14E-05 |
| ukb-b-6353 (IEU OpenGWAS) | 36 | 8.977 | 2.188, 36.829 | 2.195 | 0.720 | 2.31E-03 |
| FinnGen (R5) | 0 | - | - | - | - | - |
| FinnGen (R9) | 5 | 1.108 | 0.992, 1.238 | 0.102 | 0.057 | 7.04E-02 |
| FinnGen (R10) | 8 | 1.059 | 0.966, 1.162 | 0.058 | 0.047 | 2.22E-01 |
| Newly derived (including related individuals) | 48 | 1.119 | 1.033 to 1.213 | 0.113 | 0.041 | 5.72E-03 |
| Newly derived (unrelated individuals only) | 54 | 1.105 | 1.028 to 1.188 | 0.100 | 0.037 | 6.47E-03 |

\*Estimated effect in units of log odds ratio for POAG in myopic vs. non-myopic individuals.
Abbreviations: OR=Odds ratio; CI=Confidence interval; SE=Standard error; GWAS= Genome wide association study;

## Discussion

This work revealed that MR analyses employing different sets of publicly available GWAS summary statistics for myopia can yield contradictory findings. Our first exemplar testcase examined the relationship between years of education and myopia. Prior research using a range of methods has suggested that education is a causal risk factor for myopia.^12, 13, 43, 44^ Here, we found that researchers performing an MR analysis of education and myopia would have obtained evidence of a positive causal relationship, a null relationship or even a negative causal association, depending on the choice of GWAS summary statistics. Our second exemplar testcase examined the risk of POAG in myopic vs. non-myopic individuals. Prior research generally suggests that myopia modestly increases the risk of POAG, although the findings have not always been conclusive.^14, 45–47^ Here, we found that researchers performing an MR analysis would have obtained evidence of a large protective effect of myopia on the risk of POAG (OR < 0.1), a large increased risk (OR ≈ 9.0), or a no significant relationship, depending on the choice of summary statistics for myopia. As discussed in detail below, the reason for all these disparate findings is that publicly available summary statistics for myopia are derived from poorly designed GWAS analyses in which myopia cases and controls were often misclassified.

Myopia is a common refractive error. Among the middle and older-aged populations of Europe the prevalence of myopia is approximately 30.6%.^48^ However, the benign nature of myopia in most individuals has led to myopia being under-reported in electronic health records (EHR).^49, 50^ For example, the myopia prevalence in the FinnGen sample based on EHR information (ICD code H52.1) is approximately 1% (Table 1), yet the true prevalence of myopia in Finland is 22- 30%.^41, 42^ Work by Wittenborn and colleagues suggested that as well as myopia, several other eye conditions were markedly under-reported in an EHR system.^50^ Thus, a GWAS for myopia in the FinnGen sample using EHR-based information would analyze a sample of participants with a case-control ratio of approximately 1:100; the cases would be *bona fide* myopia cases, but 20- 30% of the controls would be myopic individuals who were misclassified. When using MR to evaluate the relationship between education and myopia, these misclassification issues with the FinnGen myopia summary statistics led to loss of statistical power, such that the causal effect of education on myopia was difficult to distinguished from a null causal effect (Table 2; Figure 1). Similarly, in the MR examining the relationship between myopia and POAG, the misclassification issues with the FinnGen myopia summary statistics led to few SNPs being available as instrumental variables, leading to limited statistical power to distinguish a causal effect from a null effect.

By contrast to the FinnGen study, the UK Biobank study performed direct assessment of refractive error using autorefraction and specifically asked participants if they were nearsighted.

However, since the ophthalmic assessment component was introduced late in the UK Biobank recruitment process, only 23% of participants underwent the eye examinations and completed the ophthalmic questionnaire.^40^ The true prevalence of myopia in these UK Biobank participants was approximately 38%.^40^ When researchers from the Jiang et al.^39^ study, the Neale lab, and the IEU OpenGWAS project classified UK Biobank participants as myopia cases or controls, the lack of ophthalmic assessment information was not taken into account; instead, the 385,000 participants who were not asked about their myopia status were all classified as non-myopic controls.

Specifically, the GWAS for myopia performed by the Neale lab and IEU OpenGWAS project used 27,000 and 37,000 correctly-classified cases, respectively, but 309,000 and 423,000 controls, respectively, of whom about 38% were misclassified. As discussed below, we found that the misclassification of controls had a greater impact on downstream MR studies for the Neale lab and OpenGWAS myopia summary statistics compared to the Jiang et al. and Zhou et al. summary statistics.

An additional concern regarding existing GWAS summary statistics for myopia is the use of a highly imbalanced case-control ratio. As mentioned above, published studies have used case-control ratios as high as of 1:100 or even 1:300 (Table 1). Logistic regression analysis using a highly imbalanced case-control ratio may result in an inflated type I error rate and inaccurate estimated effect sizes;^33^ the latter phenomenon would directly impact MR causal effect estimates. Fortunately, methods for performing case-control GWAS analyses that account for case-control imbalance have been developed, such as SAIGE,^33^ fastGWA-GLMM,^39^ GMMAT,^51^ and others.^52^ Of the sets of myopia summary statistics from publicly available databases, most used an appropriate method to address the case-control imbalance: Jiang et al.^39^ used fastGWA-GLMM, while Zhou et al.^33^ and the FinnGen team^33^ employed SAIGE. However, the Neale lab and IEU OpenGWAS project utilized approaches that did not account for case-control imbalance: Hail (https://hail.is/docs/0.2/overview/index.html) and BOLT-LMM^53^, respectively. Therefore, the published GWAS summary statistics for myopia from these two sources may include inaccurate regression coefficients for some SNPs and even include false-positive SNPs that reached the genome-wide significance threshold for spurious reasons. The combination of misclassification of controls and insufficient account of case-control imbalance is a potential reason for the worse performance of the Neale lab and IEU OpenGWAS project myopia summary statistics in our exemplar MR scenarios, compared to the summary statistics released by Jiang et al. and Zhou et al. Thus, whereas the Jiang et al. and Zhou et al. myopia summary statistics yielded MR causal effect estimates comparable to those of our gold standard summary statistics, MR causal effect estimates obtained using the Neale lab and IEU OpenGWAS project were grossly misleading in suggesting highly significant causal effects of very different magnitude from our gold standard causal effect estimates (Tables 2-3, Figures 1-2). Finally, the GWAS summary statistics from the Neale lab used in the current study had wrongly labeled risk and reference alleles, which led to MR causal effect estimates in the opposite direction to that expected, for example suggesting that additional education reduced the risk of myopia. We downloaded the Neale lab myopia summary statistics from the IEU OpenGWAS repository, since the original Neale lab GWAS repository no longer exists and because the IEU OpenGWAS repository is the most widely used source of myopia summary statistics for use in MR studies (Table 1). We were able to confirm that a more recent release of myopia summary statistics by the Neale lab has the risk and effect alleles correctly labelled (however, the misclassification of controls and the lack of account for case-control imbalance remain as potential issues).

## Conclusions

Mendelian randomization analyses performed using different sets of publicly available GWAS summary statistics for myopia can yield conflicting results. The results of previously published MR studies that have used these resources may not be valid (Table 1). Although we investigated just two exemplar scenarios – the impact of education on myopia and the risk of POAG conferred by myopia – we suspect this issue of inappropriate GWAS summary statistics may extend to other ophthalmic diseases. We urge researchers to exercise caution when selecting GWAS summary statistics for an MR study: The adage “choose the GWAS study with the largest sample size” may not always hold.^10^ Specifically, researchers should consider the underlying population, case definition, and method of association analyses when selecting summary statistics for MR. We have made our two newly derived sets of myopia summary statistics openly available and encourage researchers to utilize these for future MR studies of myopia.

## Acknowledgements

This research was conducted using the UK Biobank Resource under Application Number 83325. Data analysis was performed on the HAWK computing cluster, managed by Supercomputing Wales and Cardiff University ARCCA. We would like to thank the UK Biobank, FinnGen, and all participants who contributed to the two cohorts. Additionally, we extend our sincere gratitude to the authors of all GWAS who made their summary statistics available to facilitate this work.

## Data Availability

UK Biobank data are available via an application to the study’s access team (https://www.ukbiobank.ac.uk/enable-your-research/about-our-data). GWAS summary statistics for POAG were downloaded from the GWAS catalog (https://www.ebi.ac.uk/gwas/studies/GCST90011766). Existing GWAS summary statistics for myopia were downloaded from the GWAS catalog (Jiang et al. http://ftp.ebi.ac.uk/pub/databases/gwas/summary_statistics/GCST90044001-GCST90045000/GCST90044326 ; Zhou et al. http://ftp.ebi.ac.uk/pub/databases/gwas/summary_statistics/GCST90435001-GCST90436000/GCST90435990), the IEU OpenGWAS database (Neale lab: https://gwas.mrcieu.ac.uk/files/ukb-a-419/ukb-a-419.vcf.gz ; MRC IEU: https://gwas.mrcieu.ac.uk/files/ukb-b-6353/ukb-b-6353.vcf.gz) and the FinnGen project (R5: https://storage.googleapis.com/finngen-public-data-r5/summary_stats/finngen_R5_H7_MYOPIA.gz ; R9: https://storage.googleapis.com/finngen-public-data-r9/summary_stats/finngen_R9_H7_MYOPIA.gz ; R10: https://storage.googleapis.com/finngen-public-data-r10/summary_stats/finngen_R10_H7_MYOPIA.gz). New summary statistics from a GWAS of myopia in related and unrelated UK Biobank participants are openly available (doi: 10.17035/cardiff.29211071).

## Notes

### Competing Interest Statement

The authors have declared no competing interest.

### Funding Statement

This work was funded by UK Research and Innovation (UKRI) under the UK government's Horizon Europe funding guarantee [grant number EP/Y032292/1]. Funded by the European Union (Project 101119501. MyoTreat: HORIZON-MSCA-2022-DN-01). Views and opinions expressed are however those of the author(s) only and do not necessarily reflect those of the European Union or UKRI. Neither the European Union nor the granting authority can be held responsible for them.

### Author Declarations

Data were available before the start of the study or have been made openly available by the authors. https://research-data.cardiff.ac.uk/articles/dataset/Genome-wide_association_studies_for_myopia_in_related_and_unrelated_individuals/29211071/1

