## Supplementary Material for "Mendelian Randomization Studies of Myopia: Choosing the right Summary Statistics"

**Supplementary Note S1. Existing publicly available GWAS summary statistics for myopia**

**GCST90435990***, Zhou et al.*[1] This GWAS for myopia was reported in 2018 in an article presenting the Scalable and Accurate Implementation of Generalized mixed model (SAIGE) method. A total of 28,000,000 genetic markers were tested on a sample of 407,787 individuals from the UK Biobank using a generalized linear mixed model (GLMM)-based method. Myopia case-control status was defined using PheCode 367.1, which is based on hospital records. Since myopia is rarely listed in general medical records by the UK National Health Service, this resulted in severe misclassification bias (myopia prevalence inferred to be 0.3% rather than the true prevalence of ~38%).

**GCST90044326***, Jiang et al.*[2] This GWAS for myopia was reported in 2021 as part of an article presenting the fastGWA-GLMM method. Using a GLMM-based method, 11,842,647 SNPs were tested in a sample of 455,654 individuals from the UK Biobank. Myopia was defined based on UKB data field 6147 (Reason for glasses/contact lenses: For short-sightedness). However, Jiang et al. did not take into account that only 25% of UK Biobank participants were asked this question. Participants who were not asked the question were categorized as controls, which resulted in severe misclassification bias (myopia prevalence inferred to be 8% rather than the true prevalence of ~38%).

**ukb-b-6353**. This GWAS for myopia was conducted between 2017 and 2018 on a sample of 460,536 individuals from the UK Biobank by the MRC-IEU Consortium.[3] A total of 9,851,867 SNPs were tested using the linear mixed model association method implemented in BOLT-LMM (version 2.3). Myopia case-control status was defined based on UKB data field 6147 (Reason for glasses/contact lenses: For short-sightedness). However, the researchers did not take into account that only 25% of UK Biobank participants were asked this question. Participants who were not asked the question were categorized as controls, which resulted in severe misclassification bias (myopia prevalence inferred to be 8% rather than the true prevalence of ~38%).

**ukb-a-419**. This GWAS for myopia was conducted in 2017 in a sample of 335,700 individuals from the UK Biobank by the Neale Lab. A total of 10,894,596 SNPs were tested using the linear regression model using Hail. A comprehensive description of the pipeline is provided in their Github repository (https://github.com/Nealelab/UK_Biobank_GWAS/tree/master/imputed-v2-gwas#association-in-hail). Myopia case-control status was defined based on UKB data field 6147 (Reason for glasses/contact lenses: For short-sightedness). However, the researchers did not take into account that only 25% of UK Biobank participants were asked this question. Participants who were not asked the question were categorized as controls, which resulted in severe misclassification bias (myopia prevalence inferred to be 8% rather than the true prevalence of ~38%).

**H7_MYOPIA and finn-b-H7_MYOPIA.**[4] The FinnGen consortium have published twelve sets of GWAS summary statistic for myopia (one set for each of the twelve FinnGen data releases). The most recent public data release (R12) was in November 2024; this included 482,182 individuals in the myopia association analysis 5,406 cases and 476,776 controls). A total of 21,311,644 variants were tested using SAIGE (v.0.35.8.8). Earlier releases used in myopia MR studies included 1,640 cases and 210,931 controls (R5), 3,534 cases and 361,237 controls (R9) and 4,106 cases and 394,028 controls (R10). Myopia was defined based on the ICD code H52.1 from EHR. Since myopia is rarely listed in EHR, this resulted in severe misclassification bias (myopia prevalence inferred to be approximately 1%).

**Supplementary Note S2. Newly-performed GWAS for myopia**

*Sample selection*. We identified UK Biobank participants who underwent refractive error measurement (Tomey RC5000 autorefractor; data field #5084-5088). The SER for each eye was calculated as: spherical power + (cylindrical power * 0.5). To ensure that the GWAS sample was drawn from the same ancestry group as individuals in SSGAC and the Gharahkhani et al.[5] GWAS, we restricted the analysis to participants of European ancestry (defined as individuals whose first two genetic principal components [PCs; data field #22009] were within the mean ± 3 standard deviations of all unrelated UK Biobank participants who self-reported as White British). Additionally, individuals were required to have heterozygosity [data field #22004] within the mean ± 4 standard deviations of all unrelated participants who self-reported as White British. Subsequently, participants were excluded if their missing genotyping rate [data field #22005] exceeded 0.05 or if their genetic sex [data field #22001] did not match their self-reported sex. Lastly, we retained participants who had no history of eye surgery (cataract [data field #5324], corneal [data field #5328], strabismus [data field 6147]). This resulted in a sample of 93,041 related individuals and a sample of 66,776 unrelated individuals. The age of the participants at the assessment visit was calculated from their date of visit [data field #53] and their month and year of birth [data fields #52 and #34, respectively].

*Association analyses.* A GWAS for myopia in unrelated individuals was carried out with logistic-Firth hybrid regression model using Plink2.[6] Age, sex, genotyping array, and the first ten PCs were included as covariates. A total of 9,802,868 imputed genetic variants with minor allele frequency (MAF) > 0.01 and per variant genotyping call rate ≥ 0.9 were tested in 66,773 individuals (25,804 cases : 40,969 controls). A GWAS that included related individuals was performed using SAIGE/GATE v0.44. Age, sex, genotyping array, and the first four PCs were included as covariates, following the covariates used by Zhou et al.[1] A total of 9,850,118 imputed genetic variants with minor allele frequency MAF > 0.01 and imputation quality INFO > 0.6 were tested on 93,036 individuals (35,531 cases : 57,505 controls).

**Supplementary Note S3. Mendelian randomization analysis**

IVW-MR, MR-EGGER, weighted median MR, and mode-based MR analyses were carried out using the R package MendelianRandomization.[7] MR-PRESSO analysis was carried out using the R package MR-PRESSO.[8] The R package LDlinkR[9] was used to check for LD between variants in the summary statistics of Okbay et al.[10] with the GBR population as the LD reference panel. Details of the 62 IVs used in the education-myopia MR can be found in Supplementary Table S1. These 62 IVs were selected from the 74 genome-wide significant variants reported by Okbay et al.[10] after excluding variants in LD ($r^2 > 0.05$) and excluding variants not available in all nine sets of myopia summary statistics.

### Supplementary Note S4. Statistical code to reproduce the analyses.

```
library(LDlinkR)
library(data.table)
library(MendelianRandomization)
library(MRPRESSO)
library(plyr)
library(ggplot2)
library(cowplot)

rm(list=ls())

####

# Files
# -----

mydir                         <- [ path of directory holding data files ]

okbay_sumstats_file           <- paste0(mydir, "Okbay_SNPs_2025-03-17.csv")
okbay_ldmat_file              <- paste0(mydir, "Okbay_SNPs_LDmatrix_2025-03-17.csv")
merged_sumstats_file          <- paste0(mydir, "Okbay_Merged_2025-03-17.csv")
tableS2_file                  <- paste0(mydir, "Manuscript/JOURNAL_TableS2_2025-05-28.csv")
table2_file                   <- paste0(mydir, "Manuscript/JOURNAL_Table2_2025-05-28.csv")
tableS1_file                  <- paste0(mydir, "Manuscript/JOURNAL_TableS1_2025-05-28.csv")
final_snplist_file            <- paste0(mydir, "Okbay_MR-SNPlist_2025-05-28.csv")

# Parameters
# ----------

snp_ld_r2_threshold   <- 0.05
convert_eduyears_sd   <- 3.6
skip_mr_presso        <- FALSE

# Read in Okbay SNP sumstats and calculate SE
# ---------------------------------------------

dataOKBAY             <- read.csv(file=okbay_sumstats_file, header=TRUE)
names(dataOKBAY)      <- c("SNP","CHR","POS_GRCh37","EA","FreqEA","BETA_TEMP","P_OKBAY")
dataOKBAY$BETA_OKBAY <- dataOKBAY$BETA_TEMP*convert_eduyears_sd
dataOKBAY$Z           <- sign(dataOKBAY$BETA_OKBAY) * abs( qnorm(dataOKBAY$P_OKBAY/2))
dataOKBAY$SE_OKBAY    <- dataOKBAY$BETA_OKBAY/dataOKBAY$Z
dataOKBAY$Z           <- NULL
num_snps              <- nrow(dataOKBAY)

# Create a squared cor^2 (R2) LD matrix using LDlink
# ----------------------------------------------------

if(file.exists(okbay_ldmat_file)!=TRUE){

progress_max          <- num_snps^2
ldmatOKBAY            <- matrix(nrow=num_snps, ncol=num_snps)
rownames(ldmatOKBAY) <- dataOKBAY$SNP
colnames(ldmatOKBAY) <- dataOKBAY$SNP

for(s1 in 1:num_snps){
for(s2 in 1:num_snps){
  if(s1==s2){
  ldmatOKBAY[s1,s2]    <- 1
  } else {
  if(dataOKBAY$CHR[s1]==dataOKBAY$CHR[s2]){
     x                 <- suppressMessages(LDmatrix(snps =
c(dataOKBAY$SNP[s1],dataOKBAY$SNP[s2]), pop = "GBR", r2d = "r2", token = mytoken))
     ldmatOKBAY[s1,s2] <- ifelse(ncol(x)==3, x[1,3], NA)
     ldmatOKBAY[s2,s1] <- ifelse(ncol(x)==3, x[1,3], NA)
     } else {
     ldmatOKBAY[s1,s2] <- 0
     ldmatOKBAY[s2,s1] <- 0
     }
     }
```

```
}
}
write.csv(ldmatOKBAY, file=okbay_ldmat_file, row.names=FALSE)
}

###############################

# HARMONIZE SUMSTATS
# ------------------

# FinnGen R5

sumstats_file="finngen_R5_H7_MYOPIA.gz"
snp_file="okbay_snps74.txt"
merge_file="finngen_R5_H7_MYOPIA_okbay_snps74.txt"

# [system commands]
# cd ${mydir2}
# tail -n +2 ${okbay_file} | awk 'BEGIN {FS=","}{print $1}' > ${snp_file}
# gunzip -c ${sumstats_file} | head -1 > ${merge_file}
# gunzip -c ${sumstats_file} | grep -w -f ${snp_file} >> ${merge_file}
# sed -e 's/#//' ${merge_file} > temp.txt
# mv temp.txt ${merge_file}
# cp ${merge_file} ${mydir}${merge_file}

dataFinnGenR5            <- as.data.frame(fread(file=paste0(mydir,merge_file), header=TRUE))
dataFinnGenR5_1          <-
dataFinnGenR5[,c("rsids","chrom","pos","ref","alt","beta","sebeta","pval")]
names(dataFinnGenR5_1)   <-
c("SNP","CHR","POS_GRCh38","NEA_FinnGenR5","EA_FinnGenR5","BETA_Temp","SE_FinnGenR5","P_FinnGenR5
")
dataM                    <- merge(dataOKBAY,dataFinnGenR5_1, by=c("SNP","CHR"), all=TRUE)
dataM$BETA_FinnGenR5     <- ifelse(dataM$EA==dataM$EA_FinnGenR5, dataM$BETA_Temp, NA)
dataM$BETA_FinnGenR5     <- ifelse(dataM$EA==dataM$NEA_FinnGenR5, -1*dataM$BETA_Temp,
dataM$BETA_FinnGenR5)
dataM$NEA_FinnGenR5      <- NULL
dataM$EA_FinnGenR5       <- NULL
dataM$BETA_Temp          <- NULL
dataM$POS_GRCh38         <- NULL
write.csv(dataM, file=merged_sumstats_file, row.names=FALSE)

# FinnGen R9

sumstats_file="finngen_R9_H7_MYOPIA.gz"
snp_file="okbay_snps74.txt"
merge_file="finngen_R9_H7_MYOPIA_okbay_snps74.txt"

# [system commands]
# cd ${mydir2}
# tail -n +2 ${okbay_file} | awk 'BEGIN {FS=","}{print $1}' > ${snp_file}
# gunzip -c ${sumstats_file} | head -1 > ${merge_file}
# gunzip -c ${sumstats_file} | grep -w -f ${snp_file} >> ${merge_file}
# sed -e 's/#//' ${merge_file} > temp.txt
# mv temp.txt ${merge_file}
# cp ${merge_file} ${mydir}${merge_file}

dataOKBAY                <- as.data.frame(fread(file=merged_sumstats_file, header=TRUE))
dataFinnGenR9            <- as.data.frame(fread(file=paste0(mydir,merge_file), header=TRUE))
dataFinnGenR9_1          <-
dataFinnGenR9[,c("rsids","chrom","pos","ref","alt","beta","sebeta","pval")]
names(dataFinnGenR9_1)   <-
c("SNP","CHR","POS_GRCh38","NEA_FinnGenR9","EA_FinnGenR9","BETA_Temp","SE_FinnGenR9","P_FinnGenR9
")
dataM                    <- merge(dataOKBAY,dataFinnGenR9_1, by=c("SNP","CHR"), all=TRUE)
dataM$BETA_FinnGenR9     <- ifelse(dataM$EA==dataM$EA_FinnGenR9, dataM$BETA_Temp, NA)
dataM$BETA_FinnGenR9     <- ifelse(dataM$EA==dataM$NEA_FinnGenR9, -1*dataM$BETA_Temp,
dataM$BETA_FinnGenR9)
dataM$NEA_FinnGenR9      <- NULL
dataM$EA_FinnGenR9       <- NULL
dataM$BETA_Temp          <- NULL
dataM$POS_GRCh38         <- NULL
```

```
write.csv(dataM, file=merged_sumstats_file, row.names=FALSE)

# FinnGen R10

sumstats_file="finngen_R10_H7_MYOPIA.gz"
snp_file="okbay_snps74.txt"
merge_file="finngen_R10_H7_MYOPIA_okbay_snps74.txt"

# [system commands]
# cd ${mydir2}
# tail -n +2 ${okbay_file} | awk 'BEGIN {FS=","}{print $1}' > ${snp_file}
# gunzip -c ${sumstats_file} | head -1 > ${merge_file}
# gunzip -c ${sumstats_file} | grep -w -f ${snp_file} >> ${merge_file}
# sed -e 's/#//' ${merge_file} > temp.txt
# mv temp.txt ${merge_file}
# cp ${merge_file} ${mydir}${merge_file}

dataOKBAY                <- as.data.frame(fread(file=merged_sumstats_file, header=TRUE))
dataFinnGenR10           <- as.data.frame(fread(file=paste0(mydir,merge_file), header=TRUE))
dataFinnGenR10_1         <-
dataFinnGenR10[,c("rsids","chrom","pos","ref","alt","beta","sebeta","pval")]
names(dataFinnGenR10_1)  <-
c("SNP","CHR","POS_GRCh38","NEA_FinnGenR10","EA_FinnGenR10","BETA_Temp","SE_FinnGenR10","P_FinnGe
nR10")
dataM                    <- merge(dataOKBAY,dataFinnGenR10_1, by=c("SNP","CHR"), all=TRUE)
dataM$BETA_FinnGenR10    <- ifelse(dataM$EA==dataM$EA_FinnGenR10, dataM$BETA_Temp, NA)
dataM$BETA_FinnGenR10    <- ifelse(dataM$EA==dataM$NEA_FinnGenR10, -1*dataM$BETA_Temp,
dataM$BETA_FinnGenR10)
dataM$NEA_FinnGenR10     <- NULL
dataM$EA_FinnGenR10      <- NULL
dataM$BETA_Temp          <- NULL
dataM$POS_GRCh38         <- NULL
write.csv(dataM, file=merged_sumstats_file, row.names=FALSE)

# New PLINK GWAS in UKBB (unrelated)

sumstats_file="gwas_myopia_plink_2025-05-22.out.txt"
snp_file="okbay_snps74.txt"
merge_file="plink_unrelated_okbay_snps74.txt"

# [system commands]
# cd ${mydir2}
# tail -n +2 ${okbay_file} | awk 'BEGIN {FS=","}{print $1}' > ${snp_file}
# head -1 ${sumstats_file} > ${merge_file}
# grep -w -f ${snp_file} ${sumstats_file} >> ${merge_file}
# sed -e 's/#//' ${merge_file} > temp.txt
# mv temp.txt ${merge_file}
# cp ${merge_file} ${mydir}${merge_file}

dataOKBAY                <- as.data.frame(fread(file=merged_sumstats_file, header=TRUE))
dataPLINK                <- as.data.frame(fread(file=paste0(mydir,merge_file), header=TRUE))
dataPLINK                <- read.table(file=paste0(mydir,merge_file), header=TRUE)
dataPLINK$EA             <- dataPLINK$A1
dataPLINK$NEA            <- ifelse(dataPLINK$EA==dataPLINK$REF, dataPLINK$ALT, dataPLINK$REF)
dataPLINK_1              <- dataPLINK[,c("ID","CHROM","POS","NEA","EA","BETA","SE","p_value")]
names(dataPLINK_1 )      <-
c("SNP","CHR","POS_GRCh38","NEA_PLINK","EA_PLINK","BETA_Temp","SE_PLINK","P_PLINK")
dataM                    <- merge(dataOKBAY,dataPLINK_1, by=c("SNP","CHR"), all=TRUE)
dataM$BETA_PLINK         <- ifelse(dataM$EA==dataM$EA_PLINK, dataM$BETA_Temp, NA)
dataM$BETA_PLINK         <- ifelse(dataM$EA==dataM$NEA_PLINK, -1*dataM$BETA_Temp,
dataM$BETA_PLINK)
dataM$NEA_PLINK          <- NULL
dataM$EA_PLINK           <- NULL
dataM$BETA_Temp          <- NULL
dataM$POS_GRCh38         <- NULL
write.csv(dataM, file=merged_sumstats_file, row.names=FALSE)

# New SAIGE GWAS in UKBB (including relatives)

sumstats_file=" gwas_myopia_saige_2025-05-22.out.txt"
snp_file="okbay_snps74.txt"
```

```
merge_file="saige_related_okbay_snps74.txt"

# [system commands]
# cd ${mydir2}
# tail -n +2 ${okbay_file} | awk 'BEGIN {FS=","}{print $1}' > ${snp_file}
# head -1 ${sumstats_file} > ${merge_file}
# grep -w -f ${snp_file} ${sumstats_file} >> ${merge_file}
# sed -e 's/#//' ${merge_file} > temp.txt
# mv temp.txt ${merge_file}
# cp ${merge_file} ${mydir}${merge_file}

dataOKBAY                <- as.data.frame(fread(file=merged_sumstats_file, header=TRUE))
dataSAIGE                <- as.data.frame(fread(file=paste0(mydir,merge_file), header=TRUE))
dataSAIGE                <- read.table(file=paste0(mydir,merge_file), header=TRUE)
dataSAIGE_1              <- dataSAIGE[,c("rsid","CHR","POS","REF","ALT","BETA","SE","p.value")]
names(dataSAIGE_1 )      <-
c("SNP","CHR","POS_GRCh38","NEA_SAIGE","EA_SAIGE","BETA_Temp","SE_SAIGE","P_SAIGE")
dataM                    <- merge(dataOKBAY,dataSAIGE_1, by=c("SNP","CHR"), all=TRUE)
dataM$BETA_SAIGE         <- ifelse(dataM$EA==dataM$EA_SAIGE, dataM$BETA_Temp, NA)
dataM$BETA_SAIGE         <- ifelse(dataM$EA==dataM$NEA_SAIGE, -1*dataM$BETA_Temp,
dataM$BETA_SAIGE)
dataM$NEA_SAIGE          <- NULL
dataM$EA_SAIGE           <- NULL
dataM$BETA_Temp          <- NULL
dataM$POS_GRCh38         <- NULL
write.csv(dataM, file=merged_sumstats_file, row.names=FALSE)

# GCST90044326 (Jiang et al.)

sumstats_file="GCST90044326_buildGRCh37.tsv.gz"
snp_file="okbay_snps74.txt"
merge_file="jiang_MYOPIA_okbay_snps74.txt"

# [system commands]
# cd ${mydir2}
# tail -n +2 ${okbay_file} | awk 'BEGIN {FS=","}{print $1}' > ${snp_file}
# gunzip -c ${sumstats_file} | head -1 > ${merge_file}
# gunzip -c ${sumstats_file} | grep -w -f ${snp_file} >> ${merge_file}
# sed -e 's/#//' ${merge_file} > temp.txt
# mv temp.txt ${merge_file}
# cp ${merge_file} ${mydir}${merge_file}

dataOKBAY                <- as.data.frame(fread(file=merged_sumstats_file, header=TRUE))
dataJIANG                <- as.data.frame(fread(file=paste0(mydir,merge_file), header=TRUE))
dataJIANG_1              <-
dataJIANG[,c("variant_id","chromosome","base_pair_location","other_allele","effect_allele","beta"
,"standard_error","p_value")]
names(dataJIANG_1)       <-
c("SNP","CHR","POS_GRCh38","NEA_JIANG","EA_JIANG","BETA_Temp","SE_JIANG","P_JIANG")
dataM                    <- merge(dataOKBAY,dataJIANG_1, by=c("SNP","CHR"), all=TRUE)
dataM$BETA_JIANG         <- ifelse(dataM$EA==dataM$EA_JIANG, dataM$BETA_Temp, NA)
dataM$BETA_JIANG         <- ifelse(dataM$EA==dataM$NEA_JIANG, -1*dataM$BETA_Temp,
dataM$BETA_JIANG)
dataM$NEA_JIANG          <- NULL
dataM$EA_JIANG           <- NULL
dataM$BETA_Temp          <- NULL
dataM$POS_GRCh38         <- NULL
write.csv(dataM, file=merged_sumstats_file, row.names=FALSE)

# GCST90435990 (Zhou et al.)

sumstats_file="GCST90435990.tsv.gz"
snp_file="okbay_snps74.txt"
merge_file="zhou_MYOPIA_okbay_snps74.txt"

# [system commands]
# cd ${mydir2}
# tail -n +2 ${okbay_file} | awk 'BEGIN {FS=","}{print $1}' > ${snp_file}
# gunzip -c ${sumstats_file} | head -1 > ${merge_file}
# gunzip -c ${sumstats_file} | grep -w -f ${snp_file} >> ${merge_file}
# sed -e 's/#//' ${merge_file} > temp.txt
```

```
# mv temp.txt ${merge_file}
# cp ${merge_file} ${mydir}${merge_file}

dataOKBAY                <- as.data.frame(fread(file=merged_sumstats_file, header=TRUE))
dataZHOU                 <- as.data.frame(fread(file=paste0(mydir,merge_file), header=TRUE))
dataZHOU_1               <-
dataZHOU[,c("variant_id","chromosome","base_pair_location","other_allele","effect_allele","beta",
"standard_error","p_value")]
names(dataZHOU_1)        <-
c("SNP","CHR","POS_GRCh38","NEA_ZHOU","EA_ZHOU","BETA_Temp","SE_ZHOU","P_ZHOU")
dataM                    <- merge(dataOKBAY,dataZHOU_1, by=c("SNP","CHR"), all=TRUE)
dataM$BETA_ZHOU          <- ifelse(dataM$EA==dataM$EA_ZHOU, dataM$BETA_Temp, NA)
dataM$BETA_ZHOU          <- ifelse(dataM$EA==dataM$NEA_ZHOU, -1*dataM$BETA_Temp, dataM$BETA_ZHOU)
dataM$NEA_ZHOU           <- NULL
dataM$EA_ZHOU            <- NULL
dataM$BETA_Temp          <- NULL
dataM$POS_GRCh38         <- NULL
write.csv(dataM, file=merged_sumstats_file, row.names=FALSE)

# ukb-a-419 (Neale Lab)

sumstats_file="ukb-a-419.vcf.gz"
snp_file="okbay_snps74.txt"
merge_file="neale_MYOPIA_okbay_snps74.txt"

# [system commands]
# cd ${mydir2}
# tail -n +2 ${okbay_file} | awk 'BEGIN {FS=","}{print $1}' > ${snp_file}
# echo "SNP CHR POS REF ALT BETA SE LOG10_P" > ${merge_file}
# gunzip -c ${sumstats_file} | grep -w -f ${snp_file} > temp.txt
# awk '{split($10,a,":")}{print $3,$1,$2,$4,$5,a[1],a[2],a[3]}' temp.txt >> ${merge_file}
# sed -e 's/#//' ${merge_file} > temp.txt
# mv temp.txt ${merge_file}
# cp ${merge_file} ${mydir}${merge_file}

dataOKBAY                <- as.data.frame(fread(file=merged_sumstats_file, header=TRUE))
dataNEALE                <- as.data.frame(fread(file=paste0(mydir,merge_file), header=TRUE))
dataNEALE$P              <- 10^(-1*dataNEALE$LOG10_P)
dataNEALE_1              <- dataNEALE[,c("SNP","CHR","POS","REF","ALT","BETA","SE","P")]
names(dataNEALE_1)       <-
c("SNP","CHR","POS_GRCh38","NEA_NEALE","EA_NEALE","BETA_Temp","SE_NEALE","P_NEALE")
dataM                    <- merge(dataOKBAY,dataNEALE_1, by=c("SNP","CHR"), all=TRUE)
dataM$BETA_NEALE         <- ifelse(dataM$EA==dataM$EA_NEALE, dataM$BETA_Temp, NA)
dataM$BETA_NEALE         <- ifelse(dataM$EA==dataM$NEA_NEALE, -1*dataM$BETA_Temp,
dataM$BETA_NEALE)
dataM$NEA_NEALE          <- NULL
dataM$EA_NEALE           <- NULL
dataM$BETA_Temp          <- NULL
dataM$POS_GRCh38         <- NULL
write.csv(dataM, file=merged_sumstats_file, row.names=FALSE)

# ukb-b-6353 (MRCIEU)

sumstats_file="ukb-b-6353.vcf.gz"
snp_file="okbay_snps74.txt"
merge_file="mrcieu_MYOPIA_okbay_snps74.txt"

# [system commands]
# cd ${mydir2}
# tail -n +2 ${okbay_file} | awk 'BEGIN {FS=","}{print $1}' > ${snp_file}
# echo "SNP CHR POS REF ALT BETA SE LOG10_P" > ${merge_file}
# gunzip -c ${sumstats_file} | grep -w -f ${snp_file} > temp.txt
# awk '{split($10,a,":")}{print $3,$1,$2,$4,$5,a[1],a[2],a[3]}' temp.txt >> ${merge_file}
# sed -e 's/#//' ${merge_file} > temp.txt
# mv temp.txt ${merge_file}
# cp ${merge_file} ${mydir}${merge_file}

dataOKBAY                <- as.data.frame(fread(file=merged_sumstats_file, header=TRUE))
dataMRCIEU               <- as.data.frame(fread(file=paste0(mydir,merge_file), header=TRUE))
dataMRCIEU$P             <- 10^(-1*dataMRCIEU$LOG10_P)
```

```
dataMRCIEU_1             <- dataMRCIEU[,c("SNP","CHR","POS","REF","ALT","BETA","SE","P")]
names(dataMRCIEU_1)      <-
c("SNP","CHR","POS_GRCh38","NEA_MRCIEU","EA_MRCIEU","BETA_Temp","SE_MRCIEU","P_MRCIEU")
dataM                    <- merge(dataOKBAY,dataMRCIEU_1, by=c("SNP","CHR"), all=TRUE)
dataM$BETA_MRCIEU        <- ifelse(dataM$EA==dataM$EA_MRCIEU, dataM$BETA_Temp, NA)
dataM$BETA_MRCIEU        <- ifelse(dataM$EA==dataM$NEA_MRCIEU, -1*dataM$BETA_Temp,
dataM$BETA_MRCIEU)
dataM$NEA_MRCIEU         <- NULL
dataM$EA_MRCIEU          <- NULL
dataM$BETA_Temp          <- NULL
dataM$POS_GRCh38         <- NULL
write.csv(dataM, file=merged_sumstats_file, row.names=FALSE)

################################

# MENDELIAN RANDOMIZATION
# ----------------------

ldmatOKBAY           <- as.matrix(read.csv(file=okbay_ldmat_file, header=TRUE))
rownames(ldmatOKBAY) <- colnames(ldmatOKBAY)
num_snps             <- nrow(ldmatOKBAY)
dataT                <- as.data.frame(cbind(colnames(ldmatOKBAY),1:num_snps))
names(dataT)         <- c("SNP","SortOrder")
dataOKBAY            <- read.csv(file=merged_sumstats_file, header=TRUE)
dataOKBAY            <- merge(dataOKBAY,dataT,by="SNP")
dataOKBAY            <- dataOKBAY[order(as.numeric(dataOKBAY$SortOrder)),]

# Remove SNPs not present in all datasets

dataFULL             <- dataOKBAY[complete.cases(dataOKBAY),]
bad_snps             <- as.numeric(dataOKBAY[!dataOKBAY$SNP %in% dataFULL$SNP,]$SortOrder)
ldmatOKBAY           <- ldmatOKBAY[-bad_snps,-bad_snps]
num_snps             <- num_snps - length(bad_snps)

# Remove SNPs in LD

removed_snps         <- NULL
ldOK                 <- 0
while(ldOK==0){
bad_ld_snps          <- NULL
for (row_snp in 1:num_snps){
for (col_snp in row_snp:num_snps){
   x                 <- ifelse(is.na(ldmatOKBAY[col_snp, row_snp]),0,ldmatOKBAY[col_snp,
row_snp])
   if(x > snp_ld_r2_threshold & col_snp!=row_snp){ bad_ld_snps <- c(bad_ld_snps, col_snp,
row_snp) }
}
}
if(length(bad_ld_snps)>0){
  x1                <- as.data.frame(table(bad_ld_snps))
  x2                <- x1[order(-x1$Freq),]
  worst_snp         <- as.numeric(as.character(x2$bad_ld_snps[1]))
  removed_snps      <- c(removed_snps, colnames(ldmatOKBAY)[worst_snp])
  ldmatOKBAY        <- ldmatOKBAY[-worst_snp,-worst_snp]
  num_snps          <- num_snps - 1
  }
if(length(bad_ld_snps)==0){  ldOK=1  }
}

# Create a table for the results

fullres                           <- as.data.frame(matrix(ncol=14, nrow=2))
names(fullres)                    <-
c("Method","Sumstats","Num_IVs","OR","LCI95","UCI95","BETA","SE","P","Qstat",
"Qpval","Egger_intercept","Egger_P","MRP_outliers")
myrow                             <- 1

# Run MR
```

```
sumstats_list       <- 
c("JIANG","ZHOU","NEALE","MRCIEU","FinnGenR5","FinnGenR9","FinnGenR10","SAIGE","PLINK")
dataMR              <- dataFULL[!dataFULL$SNP %in% removed_snps,]

for(k in 1:length(sumstats_list)){
mystats             <- sumstats_list[k]
mybeta              <- paste0("BETA_",mystats)
myse                <- paste0("SE_",mystats)
dataMR$BETA_OUT     <- dataMR[,mybeta]
dataMR$SE_OUT       <- dataMR[,myse]
MR_obj              <- MendelianRandomization::mr_input(exposure    = "EduYears",
                                                        outcome     = "Myopia status",
                                                        snps        = dataMR$SNP,
                                                        bx          = dataMR$BETA_OKBAY,
                                                        bxse        = dataMR$SE_OKBAY,
                                                        by          = dataMR[,mybeta],
                                                        byse        = dataMR[,myse])
MR_ivw               <- MendelianRandomization::mr_ivw    (MR_obj, robust = FALSE, distribution = 
"normal", penalized = FALSE, alpha = 0.05, correl = FALSE)
fullres[myrow,1]     <- "IVW-MR"
fullres[myrow,2]     <- mystats
fullres[myrow,3]     <- MR_ivw$SNPs
fullres[myrow,4]     <- sprintf("%.3f", exp(MR_ivw$Estimate))
fullres[myrow,5]     <- sprintf("%.3f", exp(MR_ivw$CILower))
fullres[myrow,6]     <- sprintf("%.3f", exp(MR_ivw$CIUpper))
fullres[myrow,7]     <- sprintf("%.3f", MR_ivw$Estimate)
fullres[myrow,8]     <- sprintf("%.3f", MR_ivw$StdError)
fullres[myrow,9]     <- sprintf("%.2e", MR_ivw$Pvalue)
fullres[myrow,10]    <- sprintf("%.3f", MR_ivw$Heter.Stat[1])
fullres[myrow,11]    <- sprintf("%.3f", MR_ivw$Heter.Stat[2])
fullres[myrow,12:14] <- c("-","-","-")
myrow                <- myrow + 1

MR_egg               <- MendelianRandomization::mr_egger    (MR_obj, robust = FALSE, distribution 
= "normal", penalized = FALSE, alpha = 0.05, correl = FALSE)
fullres[myrow,1]     <- "MR-EGGER"
fullres[myrow,2]     <- mystats
fullres[myrow,3]     <- MR_egg$SNPs
fullres[myrow,4]     <- sprintf("%.3f", exp(MR_egg$Estimate))
fullres[myrow,5]     <- sprintf("%.3f", exp(MR_egg$CILower.Est))
fullres[myrow,6]     <- sprintf("%.3f", exp(MR_egg$CIUpper.Est))
fullres[myrow,7]     <- sprintf("%.3f", MR_egg$Estimate)
fullres[myrow,8]     <- sprintf("%.3f", MR_egg$StdError.Est)
fullres[myrow,9]     <- sprintf("%.2e", MR_egg$Pvalue.Est)
fullres[myrow,10]    <- sprintf("%.3f", MR_egg$Heter.Stat[1])
fullres[myrow,11]    <- sprintf("%.3f", MR_egg$Heter.Stat[2])
fullres[myrow,12]    <- sprintf("%.3f", MR_egg$Intercept)
fullres[myrow,13]    <- sprintf("%.2e", MR_egg$Pvalue.Int)
fullres[myrow,14]    <- "-"
myrow                <- myrow + 1

MR_median            <- MendelianRandomization::mr_median    (MR_obj, distribution = "normal", 
weighting = "weighted", alpha = 0.05)
fullres[myrow,1]     <- "MR-WMEDIAN"
fullres[myrow,2]     <- mystats
fullres[myrow,3]     <- MR_median$SNPs
fullres[myrow,4]     <- sprintf("%.3f", exp(MR_median$Estimate))
fullres[myrow,5]     <- sprintf("%.3f", exp(MR_median$CILower))
fullres[myrow,6]     <- sprintf("%.3f", exp(MR_median$CIUpper))
fullres[myrow,7]     <- sprintf("%.3f", MR_median$Estimate)
fullres[myrow,8]     <- sprintf("%.3f", MR_median$StdError)
fullres[myrow,9]     <- sprintf("%.2e", MR_median$Pvalue)
fullres[myrow,10:14] <- c("-","-","-","-","-")
myrow                <- myrow + 1

MR_mbe               <- MendelianRandomization::mr_mbe    (MR_obj, distribution = "normal", 
weighting = "weighted", stderror = "simple", phi = 1, alpha = 0.05)
fullres[myrow,1]     <- "MR-MBE"
fullres[myrow,2]     <- mystats
fullres[myrow,3]     <- MR_mbe$SNPs
fullres[myrow,4]     <- sprintf("%.3f", exp(MR_mbe$Estimate))
```

```
fullres[myrow,5]      <- sprintf("%.3f", exp(MR_mbe$CILower))
fullres[myrow,6]      <- sprintf("%.3f", exp(MR_mbe$CIUpper))
fullres[myrow,7]      <- sprintf("%.3f", MR_mbe$Estimate)
fullres[myrow,8]      <- sprintf("%.3f", MR_mbe$StdError)
fullres[myrow,9]      <- sprintf("%.2e", MR_mbe$Pvalue)
fullres[myrow,10:14] <- c("-","-","-","-","-")
myrow                 <- myrow + 1

if(!skip_mr_presso){
 MR_mrp               <-  mr_presso(BetaOutcome = "BETA_OUT", BetaExposure = "BETA_OKBAY",
SdOutcome = "SE_OUT", SdExposure = "SE_OKBAY",
                           OUTLIERtest = TRUE, DISTORTIONtest = TRUE, NbDistribution = 1000,
SignifThreshold = 0.05, data=dataMR)

 num_ouliers          <- sum(MR_mrp$MR$Outlier[,2]<0.05)
 h                    <- ifelse(num_ouliers>0, 2, 1)
 LCI                  <- MR_mrp[[1]]$Causal[h] - (1.96*MR_mrp[[1]]$Sd[h])
 UCI                  <- MR_mrp[[1]]$Causal[h] + (1.96*MR_mrp[[1]]$Sd[h])
 fullres[myrow,1]     <- "MR-PRESSO"
 fullres[myrow,2]     <- mystats
 fullres[myrow,3]     <- MR_ivw$SNPs - num_ouliers
 fullres[myrow,4]     <- sprintf("%.3f", exp(MR_mrp[[1]]$Causal[h]))
 fullres[myrow,5]     <- sprintf("%.3f", exp(LCI))
 fullres[myrow,6]     <- sprintf("%.3f", exp(UCI))
 fullres[myrow,7]     <- sprintf("%.3f", MR_mrp[[1]]$Causal[h])
 fullres[myrow,8]     <- sprintf("%.3f", MR_mrp[[1]]$Sd[h])
 fullres[myrow,9]     <- sprintf("%.2e", MR_mrp[[1]]$P[h])
 fullres[myrow,10:13] <- c("-","-","-","-")
 fullres[myrow,14]    <- num_ouliers
 myrow                <- myrow + 1
 }
}
fullres
ivw_res                  <- fullres[which(fullres$Method=="IVW-MR"),]
ivw_res$ci95             <- paste0(ivw_res$LCI95, " to ", ivw_res$UCI95)
write.csv(fullres, file=tableS2_file, row.names=FALSE)
write.csv(ivw_res[,c("Sumstats","OR","ci95","BETA","SE","P")], file=table2_file, row.names=FALSE)
write.csv(dataMR[,c("SNP","CHR","POS_GRCh37","EA","BETA_OKBAY","SE_OKBAY","P_OKBAY")],file=tableS
1_file, row.names=FALSE)
```

---

```
# Files
# -----

poag_sumstats_file         <- paste0(mydir, "clumped/poag_sumstats_for_clumped_vars_2025-05-
20.txt")
merged_sumstats_file       <- paste0(mydir, "poag_merged_sumstats_2025-05-20.txt")
tableS3_file               <- paste0(mydir, "Manuscript/JOURNAL_TableS3_2025-05-28.csv")
table3_file                <- paste0(mydir, "Manuscript/JOURNAL_Table3_2025-05-28.csv")
clumped_zhou_file          <- paste0(mydir, "clumped/zhou_2025-05-20_clumped.out")
clumped_jiang_file         <- paste0(mydir, "clumped/jiang_2025-05-20_clumped.out")
clumped_neale_file         <- paste0(mydir, "clumped/neale419_2025-05-20_clumped.out")
clumped_mrcieu_file        <- paste0(mydir, "clumped/mrcieu6353_2025-05-20_clumped.out")
clumped_finngenR9_file     <- paste0(mydir, "clumped/finngen_r9_2025-05-20_clumped.out")
clumped_finngenR10_file    <- paste0(mydir, "clumped/finngen_r10_2025-05-20_clumped.out")
clumped_new_plink_file     <- paste0(mydir, "clumped/plink_new_2025-05-20_clumped.out")
clumped_new_saige_file     <- paste0(mydir, "clumped/saige_new_2025-05-20_clumped.out")

# Parameters
# ----------

skip_mr_presso             <- FALSE

###############################

# HARMONIZE SUMSTATS
# ------------------

# SAIGE/GATE
```

```
dataPOAG                 <- as.data.frame(fread(file=poag_sumstats_file, header=TRUE))
dataSAIGE                <- as.data.frame(fread(file=clumped_new_saige_file, header=TRUE))

names(dataSAIGE)         <-
c("SNP","CHR","POS_SAIGE","EA_SAIGE","NEA_SAIGE","BETA_Temp","SE_SAIGE","P_SAIGE")
names(dataPOAG)          <-
c("CHR","POS","SNP","EA_POAG","NEA_POAG","BETA_POAG","SE_POAG","P_POAG")
dataM                    <- merge(dataPOAG,dataSAIGE, by=c("SNP","CHR"), all=TRUE)
dataM$BETA_SAIGE         <- ifelse(dataM$EA_POAG==dataM$EA_SAIGE, dataM$BETA_Temp, NA)
dataM$BETA_SAIGE         <- ifelse(dataM$EA_POAG==dataM$NEA_SAIGE, -1*dataM$BETA_Temp,
dataM$BETA_SAIGE)
dataM$NEA_SAIGE          <- NULL
dataM$EA_SAIGE           <- NULL
dataM$BETA_Temp          <- NULL
dataM$POS_SAIGE          <- NULL
write.csv(dataM, file=merged_sumstats_file, row.names=FALSE)

# PLINK

dataPOAG                 <- as.data.frame(fread(file=merged_sumstats_file, header=TRUE))
dataPLINK                <- as.data.frame(fread(file=clumped_new_plink_file, header=TRUE))

names(dataPLINK)         <-
c("SNP","CHR","POS_PLINK","EA_PLINK","NEA_PLINK","BETA_Temp","SE_PLINK","P_PLINK")
dataM                    <- merge(dataPOAG,dataPLINK, by=c("SNP","CHR"), all=TRUE)
dataM$BETA_PLINK         <- ifelse(dataM$EA_POAG==dataM$EA_PLINK, dataM$BETA_Temp, NA)
dataM$BETA_PLINK         <- ifelse(dataM$EA_POAG==dataM$NEA_PLINK, -1*dataM$BETA_Temp,
dataM$BETA_PLINK)
dataM$NEA_PLINK          <- NULL
dataM$EA_PLINK           <- NULL
dataM$BETA_Temp          <- NULL
dataM$POS_PLINK          <- NULL
write.csv(dataM, file=merged_sumstats_file, row.names=FALSE)

# ZHOU

dataPOAG                 <- as.data.frame(fread(file=merged_sumstats_file, header=TRUE))
dataZHOU                 <- as.data.frame(fread(file=clumped_zhou_file, header=TRUE))

names(dataZHOU)          <-
c("SNP","CHR","POS_ZHOU","EA_ZHOU","NEA_ZHOU","BETA_Temp","SE_ZHOU","P_ZHOU")
dataM                    <- merge(dataPOAG,dataZHOU, by=c("SNP","CHR"), all=TRUE)
dataM$BETA_ZHOU          <- ifelse(dataM$EA_POAG==dataM$EA_ZHOU, dataM$BETA_Temp, NA)
dataM$BETA_ZHOU          <- ifelse(dataM$EA_POAG==dataM$NEA_ZHOU, -1*dataM$BETA_Temp,
dataM$BETA_ZHOU)
dataM$NEA_ZHOU           <- NULL
dataM$EA_ZHOU            <- NULL
dataM$BETA_Temp          <- NULL
dataM$POS_ZHOU           <- NULL
write.csv(dataM, file=merged_sumstats_file, row.names=FALSE)

# JIANG

dataPOAG                 <- as.data.frame(fread(file=merged_sumstats_file, header=TRUE))
dataJIANG                <- as.data.frame(fread(file=clumped_jiang_file, header=TRUE))

names(dataJIANG)         <-
c("SNP","CHR","POS_JIANG","EA_JIANG","NEA_JIANG","BETA_Temp","SE_JIANG","P_JIANG")
dataM                    <- merge(dataPOAG,dataJIANG, by=c("SNP","CHR"), all=TRUE)
dataM$BETA_JIANG         <- ifelse(dataM$EA_POAG==dataM$EA_JIANG, dataM$BETA_Temp, NA)
dataM$BETA_JIANG         <- ifelse(dataM$EA_POAG==dataM$NEA_JIANG, -1*dataM$BETA_Temp,
dataM$BETA_JIANG)
dataM$NEA_JIANG          <- NULL
dataM$EA_JIANG           <- NULL
dataM$BETA_Temp          <- NULL
dataM$POS_JIANG          <- NULL
write.csv(dataM, file=merged_sumstats_file, row.names=FALSE)

# MRCIEU
```

```
dataPOAG                  <- as.data.frame(fread(file=merged_sumstats_file, header=TRUE))
dataMRCIEU                <- as.data.frame(fread(file=clumped_mrcieu_file, header=TRUE))

names(dataMRCIEU)         <-
c("SNP","CHR","POS_MRCIEU","EA_MRCIEU","NEA_MRCIEU","BETA_Temp","SE_MRCIEU","P_MRCIEU")
dataM                     <- merge(dataPOAG,dataMRCIEU, by=c("SNP","CHR"), all=TRUE)
dataM$BETA_MRCIEU         <- ifelse(dataM$EA_POAG==dataM$EA_MRCIEU, dataM$BETA_Temp, NA)
dataM$BETA_MRCIEU         <- ifelse(dataM$EA_POAG==dataM$NEA_MRCIEU, -1*dataM$BETA_Temp,
dataM$BETA_MRCIEU)
dataM$NEA_MRCIEU          <- NULL
dataM$EA_MRCIEU           <- NULL
dataM$BETA_Temp           <- NULL
dataM$POS_MRCIEU          <- NULL
write.csv(dataM, file=merged_sumstats_file, row.names=FALSE)

# NEALE

dataPOAG                  <- as.data.frame(fread(file=merged_sumstats_file, header=TRUE))
dataNEALE                 <- as.data.frame(fread(file=clumped_neale_file, header=TRUE))

names(dataNEALE)          <-
c("SNP","CHR","POS_NEALE","EA_NEALE","NEA_NEALE","BETA_Temp","SE_NEALE","P_NEALE")
dataM                     <- merge(dataPOAG,dataNEALE, by=c("SNP","CHR"), all=TRUE)
dataM$BETA_NEALE          <- ifelse(dataM$EA_POAG==dataM$EA_NEALE, dataM$BETA_Temp, NA)
dataM$BETA_NEALE          <- ifelse(dataM$EA_POAG==dataM$NEA_NEALE, -1*dataM$BETA_Temp,
dataM$BETA_NEALE)
dataM$NEA_NEALE           <- NULL
dataM$EA_NEALE            <- NULL
dataM$BETA_Temp           <- NULL
dataM$POS_NEALE           <- NULL
write.csv(dataM, file=merged_sumstats_file, row.names=FALSE)

# FinnGenR9

dataPOAG                  <- as.data.frame(fread(file=merged_sumstats_file, header=TRUE))
dataFinnGenR9             <- as.data.frame(fread(file=clumped_finngenR9_file, header=TRUE))

names(dataFinnGenR9)      <-
c("SNP","CHR","POS_FinnGenR9","EA_FinnGenR9","NEA_FinnGenR9","BETA_Temp","SE_FinnGenR9","P_FinnGe
nR9")
dataM                     <- merge(dataPOAG,dataFinnGenR9, by=c("SNP","CHR"), all=TRUE)
dataM$BETA_FinnGenR9      <- ifelse(dataM$EA_POAG==dataM$EA_FinnGenR9, dataM$BETA_Temp, NA)
dataM$BETA_FinnGenR9      <- ifelse(dataM$EA_POAG==dataM$NEA_FinnGenR9, -1*dataM$BETA_Temp,
dataM$BETA_FinnGenR9)
dataM$NEA_FinnGenR9       <- NULL
dataM$EA_FinnGenR9        <- NULL
dataM$BETA_Temp           <- NULL
dataM$POS_FinnGenR9       <- NULL
write.csv(dataM, file=merged_sumstats_file, row.names=FALSE)

# FinnGenR10

dataPOAG                  <- as.data.frame(fread(file=merged_sumstats_file, header=TRUE))
dataFinnGenR10            <- as.data.frame(fread(file=clumped_finngenR10_file, header=TRUE))

names(dataFinnGenR10)     <-
c("SNP","CHR","POS_FinnGenR10","EA_FinnGenR10","NEA_FinnGenR10","BETA_Temp","SE_FinnGenR10","P_Fi
nnGenR10")
dataM                     <- merge(dataPOAG,dataFinnGenR10, by=c("SNP","CHR"), all=TRUE)
dataM$BETA_FinnGenR10     <- ifelse(dataM$EA_POAG==dataM$EA_FinnGenR10, dataM$BETA_Temp, NA)
dataM$BETA_FinnGenR10     <- ifelse(dataM$EA_POAG==dataM$NEA_FinnGenR10, -1*dataM$BETA_Temp,
dataM$BETA_FinnGenR10)
dataM$NEA_FinnGenR10      <- NULL
dataM$EA_FinnGenR10       <- NULL
dataM$BETA_Temp           <- NULL
dataM$POS_FinnGenR10      <- NULL
write.csv(dataM, file=merged_sumstats_file, row.names=FALSE)

###############################
```

```
# MENDELIAN RANDOMIZATION
# -----------------------

dataM               <- read.csv(file=merged_sumstats_file, header=TRUE)

# Create a table for the results

fullres             <- as.data.frame(matrix(ncol=14, nrow=2))
names(fullres)      <-
c("Method","Sumstats","Num_IVs","OR","LCI95","UCI95","BETA","SE","P","Qstat",
"Qpval","Egger_intercept","Egger_P","MRP_outliers")
myrow               <- 1

# Run MR

sumstats_list       <-
c("JIANG","ZHOU","NEALE","MRCIEU","FinnGenR9","FinnGenR10","SAIGE","PLINK")

for(k in 1:length(sumstats_list)){
mystats             <- sumstats_list[k]
mybeta              <- paste0("BETA_",mystats)
myse                <- paste0("SE_",mystats)
dataMR              <- dataM[which(!is.na(dataM[,mybeta])),]
dataMR$BETA_EXP     <- dataMR[,mybeta]
dataMR$SE_EXP       <- dataMR[,myse]
MR_obj              <- MendelianRandomization::mr_input(exposure    = "Myopia status",
                                                        outcome     = "POAG status",
                                                        snps        = dataMR$SNP,
                                                        bx          = dataMR[,mybeta],
                                                        bxse        = dataMR[,myse],
                                                        by          = dataMR$BETA_POAG,
                                                        byse        = dataMR$SE_POAG)
MR_ivw               <- MendelianRandomization::mr_ivw    (MR_obj, robust = FALSE, distribution =
"normal", penalized = FALSE, alpha = 0.05, correl = FALSE)
fullres[myrow,1]     <- "IVW-MR"
fullres[myrow,2]     <- mystats
fullres[myrow,3]     <- MR_ivw$SNPs
fullres[myrow,4]     <- sprintf("%.3f", exp(MR_ivw$Estimate))
fullres[myrow,5]     <- sprintf("%.3f", exp(MR_ivw$CILower))
fullres[myrow,6]     <- sprintf("%.3f", exp(MR_ivw$CIUpper))
fullres[myrow,7]     <- sprintf("%.3f", MR_ivw$Estimate)
fullres[myrow,8]     <- sprintf("%.3f", MR_ivw$StdError)
fullres[myrow,9]     <- sprintf("%.2e", MR_ivw$Pvalue)
fullres[myrow,10]    <- sprintf("%.3f", MR_ivw$Heter.Stat[1])
fullres[myrow,11]    <- sprintf("%.3f", MR_ivw$Heter.Stat[2])
fullres[myrow,12:14] <- c("-","-","-")
myrow                <- myrow + 1

fullres[myrow,1]     <- "MR-EGGER"
if(nrow(dataMR)>1){
MR_egg               <- MendelianRandomization::mr_egger    (MR_obj, robust = FALSE, distribution
= "normal", penalized = FALSE, alpha = 0.05, correl = FALSE)
fullres[myrow,2]     <- mystats
fullres[myrow,3]     <- MR_egg$SNPs
fullres[myrow,4]     <- sprintf("%.3f", exp(MR_egg$Estimate))
fullres[myrow,5]     <- sprintf("%.3f", exp(MR_egg$CILower.Est))
fullres[myrow,6]     <- sprintf("%.3f", exp(MR_egg$CIUpper.Est))
fullres[myrow,7]     <- sprintf("%.3f", MR_egg$Estimate)
fullres[myrow,8]     <- sprintf("%.3f", MR_egg$StdError.Est)
fullres[myrow,9]     <- sprintf("%.2e", MR_egg$Pvalue.Est)
fullres[myrow,10]    <- sprintf("%.3f", MR_egg$Heter.Stat[1])
fullres[myrow,11]    <- sprintf("%.3f", MR_egg$Heter.Stat[2])
fullres[myrow,12]    <- sprintf("%.3f", MR_egg$Intercept)
fullres[myrow,13]    <- sprintf("%.2e", MR_egg$Pvalue.Int)
fullres[myrow,14]    <- "-"
}
myrow                <- myrow + 1

fullres[myrow,1]     <- "MR-WMEDIAN"
MR_median            <- MendelianRandomization::mr_median    (MR_obj, distribution = "normal",
weighting = "weighted", alpha = 0.05)
```

```
if(nrow(dataMR)>1){
fullres[myrow,2]     <- mystats
fullres[myrow,3]     <- MR_median$SNPs
fullres[myrow,4]     <- sprintf("%.3f", exp(MR_median$Estimate))
fullres[myrow,5]     <- sprintf("%.3f", exp(MR_median$CILower))
fullres[myrow,6]     <- sprintf("%.3f", exp(MR_median$CIUpper))
fullres[myrow,7]     <- sprintf("%.3f", MR_median$Estimate)
fullres[myrow,8]     <- sprintf("%.3f", MR_median$StdError)
fullres[myrow,9]     <- sprintf("%.2e", MR_median$Pvalue)
fullres[myrow,10:14] <- c("-","-","-","-","-")
}
myrow                <- myrow + 1

fullres[myrow,1]     <- "MR-MBE"
if(nrow(dataMR)>1){
MR_mbe               <- MendelianRandomization::mr_mbe    (MR_obj, distribution = "normal", 
weighting = "weighted", stderror = "simple", phi = 1, alpha = 0.05)
fullres[myrow,2]     <- mystats
fullres[myrow,3]     <- MR_mbe$SNPs
fullres[myrow,4]     <- sprintf("%.3f", exp(MR_mbe$Estimate))
fullres[myrow,5]     <- sprintf("%.3f", exp(MR_mbe$CILower))
fullres[myrow,6]     <- sprintf("%.3f", exp(MR_mbe$CIUpper))
fullres[myrow,7]     <- sprintf("%.3f", MR_mbe$Estimate)
fullres[myrow,8]     <- sprintf("%.3f", MR_mbe$StdError)
fullres[myrow,9]     <- sprintf("%.2e", MR_mbe$Pvalue)
fullres[myrow,10:14] <- c("-","-","-","-","-")
}
myrow                <- myrow + 1

if(!skip_mr_presso){
if(nrow(dataMR)>1){
 MR_mrp               <-  mr_presso(BetaOutcome = "BETA_POAG", BetaExposure = "BETA_EXP", 
SdOutcome = "SE_POAG", SdExposure = "SE_EXP",
                           OUTLIERtest = TRUE, DISTORTIONtest = TRUE, NbDistribution = 1000, 
SignifThreshold = 0.05, data=dataMR)

 num_ouliers          <- sum(MR_mrp$MR$Outlier[,2]<0.05)
 h                    <- ifelse(num_ouliers>0, 2, 1)
 LCI                  <- MR_mrp[[1]]$Causal[h] - (1.96*MR_mrp[[1]]$Sd[h])
 UCI                  <- MR_mrp[[1]]$Causal[h] + (1.96*MR_mrp[[1]]$Sd[h])
 fullres[myrow,1]     <- "MR-PRESSO"
 fullres[myrow,2]     <- mystats
 fullres[myrow,3]     <- MR_ivw$SNPs - num_ouliers
 fullres[myrow,4]     <- sprintf("%.3f", exp(MR_mrp[[1]]$Causal[h]))
 fullres[myrow,5]     <- sprintf("%.3f", exp(LCI))
 fullres[myrow,6]     <- sprintf("%.3f", exp(UCI))
 fullres[myrow,7]     <- sprintf("%.3f", MR_mrp[[1]]$Causal[h])
 fullres[myrow,8]     <- sprintf("%.3f", MR_mrp[[1]]$Sd[h])
 fullres[myrow,9]     <- sprintf("%.2e", MR_mrp[[1]]$P[h])
 fullres[myrow,10:13] <- c("-","-","-","-")
 fullres[myrow,14]    <- num_ouliers
 myrow                <- myrow + 1
 }
 }
}
fullres
ivw_res                  <- fullres[which(fullres$Method=="IVW-MR"),]
ivw_res$ci95             <- paste0(ivw_res$LCI95, " to ", ivw_res$UCI95)
write.csv(fullres, file=tableS3_file, row.names=FALSE)
write.csv(ivw_res[,c("Sumstats","Num_IVs","OR","ci95","BETA","SE","P")], file=table3_file, 
row.names=FALSE)

##########################
```

**Supplementary Table S1. Genetic variants (N=62) from the SSGAC GWAS for EduYears used as instrumental variables for MR.**

| SNP | CHR | POS (GRCh37) | Effect Allele | BETA | SE | *P* |
|---|---|---|---|---|---|---|
| rs301800 | 1 | 8490603 | T | 0.068 | 0.012 | 1.79e-08 |
| rs11210860 | 1 | 43982527 | A | 0.061 | 0.010 | 2.36e-10 |
| rs34305371 | 1 | 72733610 | A | 0.126 | 0.017 | 3.76e-14 |
| rs1008078 | 1 | 91189731 | T | -0.058 | 0.009 | 6.01e-10 |
| rs11588857 | 1 | 204587047 | A | 0.072 | 0.012 | 5.27e-10 |
| rs1777827 | 1 | 211613114 | A | 0.054 | 0.010 | 1.55e-08 |
| rs2992632 | 1 | 243503764 | A | 0.061 | 0.011 | 8.23e-09 |
| rs76076331 | 2 | 10977585 | T | 0.072 | 0.013 | 3.63e-08 |
| rs11689269 | 2 | 15621917 | C | 0.058 | 0.010 | 1.28e-08 |
| rs1606974 | 2 | 51873599 | A | 0.079 | 0.014 | 2.8e-08 |
| rs11690172 | 2 | 57387094 | A | 0.054 | 0.010 | 1.99e-08 |
| rs2457660 | 2 | 60757419 | T | -0.061 | 0.010 | 7.11e-10 |
| rs10496091 | 2 | 61482261 | A | -0.065 | 0.010 | 5.62e-10 |
| rs13402908 | 2 | 100333377 | T | -0.065 | 0.010 | 1.7e-11 |
| rs4851251 | 2 | 100753490 | T | -0.061 | 0.011 | 1.91e-08 |
| rs17824247 | 2 | 144152539 | T | -0.058 | 0.010 | 2.77e-09 |
| rs16845580 | 2 | 161920884 | T | 0.058 | 0.010 | 2.65e-09 |
| rs4500960 | 2 | 162818621 | T | -0.058 | 0.009 | 3.75e-10 |
| rs2245901 | 2 | 194296294 | A | -0.058 | 0.010 | 4.54e-09 |
| rs55830725 | 2 | 237056854 | A | -0.079 | 0.013 | 5.37e-10 |
| rs35761247 | 3 | 48623124 | A | 0.122 | 0.022 | 3.82e-08 |
| rs62259535 | 3 | 48939052 | A | 0.173 | 0.029 | 2.63e-09 |
| rs112634398 | 3 | 50075494 | A | 0.130 | 0.024 | 4.61e-08 |
| rs62263923 | 3 | 85674790 | A | -0.058 | 0.010 | 7.01e-09 |
| rs6799130 | 3 | 160847801 | C | -0.054 | 0.010 | 2.82e-08 |
| rs12646808 | 4 | 3249828 | T | 0.058 | 0.010 | 4e-08 |
| rs34072092 | 4 | 28801221 | T | 0.086 | 0.016 | 3.91e-08 |
| rs3101246 | 4 | 42649935 | T | -0.054 | 0.010 | 1.43e-08 |
| rs4863692 | 4 | 140764124 | T | 0.065 | 0.010 | 1.56e-10 |
| rs4493682 | 5 | 45188024 | C | 0.068 | 0.012 | 3.32e-08 |
| rs2964197 | 5 | 57535206 | T | 0.054 | 0.010 | 3.02e-08 |
| rs61160187 | 5 | 60111579 | A | -0.061 | 0.010 | 3.49e-10 |
| rs10061788 | 5 | 87934707 | A | 0.076 | 0.013 | 2.46e-09 |
| rs2431108 | 5 | 103947968 | T | 0.058 | 0.010 | 5.27e-09 |
| rs1402025 | 5 | 113987898 | T | 0.061 | 0.011 | 3.42e-08 |
| rs62379838 | 5 | 120102028 | T | 0.058 | 0.010 | 3.3e-08 |
| rs56231335 | 6 | 98187291 | T | -0.061 | 0.010 | 2.07e-09 |

| | | | | | | |
|---|---|---|---|---|---|---|
| rs7767938 | 6 | 153367613 | T | 0.061 | 0.011 | 2.44e-08 |
| rs2615691 | 7 | 23402104 | A | -0.133 | 0.024 | 4.71e-08 |
| rs12531458 | 7 | 39090698 | A | 0.050 | 0.009 | 3.11e-08 |
| rs12671937 | 7 | 92654365 | A | 0.058 | 0.009 | 9.15e-10 |
| rs113520408 | 7 | 128402782 | A | 0.061 | 0.011 | 1.97e-08 |
| rs17167170 | 7 | 133302345 | A | 0.072 | 0.012 | 1.14e-09 |
| rs11768238 | 7 | 135227513 | A | -0.061 | 0.010 | 9.9e-10 |
| rs12682297 | 8 | 145712860 | A | -0.058 | 0.010 | 3.93e-09 |
| rs1871109 | 9 | 1746016 | T | -0.058 | 0.009 | 4.35e-10 |
| rs13294439 | 9 | 23358875 | A | -0.083 | 0.010 | 2.2e-17 |
| rs895606 | 9 | 88003668 | A | 0.054 | 0.010 | 2.25e-08 |
| rs7854982 | 9 | 124644562 | T | -0.054 | 0.009 | 1.29e-08 |
| rs11191193 | 10 | 103802408 | A | 0.065 | 0.010 | 5.44e-11 |
| rs12772375 | 10 | 104082688 | T | -0.054 | 0.010 | 1.56e-08 |
| rs7945718 | 11 | 12748819 | A | 0.054 | 0.010 | 1.54e-08 |
| rs7955289 | 12 | 14653667 | A | 0.061 | 0.010 | 4.49e-10 |
| rs2456973 | 12 | 56416928 | A | -0.072 | 0.010 | 1.06e-12 |
| rs7131944 | 12 | 92159557 | A | 0.054 | 0.009 | 9.02e-09 |
| rs572016 | 12 | 121279083 | A | 0.050 | 0.009 | 3.46e-08 |
| rs9537821 | 13 | 58402771 | A | 0.086 | 0.010 | 1.5e-16 |
| rs1043209 | 14 | 23373986 | A | 0.065 | 0.010 | 1.82e-11 |
| rs17119973 | 14 | 84913111 | A | -0.068 | 0.011 | 3.55e-10 |
| rs12969294 | 18 | 35186122 | A | -0.058 | 0.010 | 7.24e-09 |
| rs2837992 | 21 | 42620520 | T | 0.054 | 0.010 | 3.8e-08 |
| rs165633 | 22 | 29880773 | A | -0.065 | 0.011 | 2.86e-09 |

**Supplementary Table S2. Full MR results for analyses examining the effect of EduYears on myopia.**

| Method | Sum. stats | Num. IVs | OR | LCI95 | UCI95 | BETA | SE | P | Qstat | Qpval | Egger intercept | Egger P | Num. MRP outliers |
|---|---|---|---|---|---|---|---|---|---|---|---|---|---|
| IVW-MR | JIANG | 62 | 1.152 | 1.097 | 1.210 | 0.141 | 0.025 | 1.63e-08 | 125.895 | 0 | - | - | - |
| MR-EGGER | JIANG | 62 | 1.010 | 0.795 | 1.284 | 0.010 | 0.122 | 9.33e-01 | 123.424 | 0 | 0.009 | 2.73e-01 | - |
| MR-WMEDIAN | JIANG | 62 | 1.174 | 1.112 | 1.240 | 0.160 | 0.028 | 8.53e-09 | - | - | - | - | - |
| MR-MBE | JIANG | 62 | 1.194 | 1.032 | 1.382 | 0.177 | 0.074 | 1.70e-02 | - | - | - | - | - |
| MR-PRESSO | JIANG | 61 | 1.162 | 1.108 | 1.219 | 0.150 | 0.024 | 5.70e-08 | - | - | - | - | 1 |
| IVW-MR | ZHOU | 62 | 1.242 | 1.043 | 1.479 | 0.217 | 0.089 | 1.49e-02 | 47.718 | 0.893 | - | - | - |
| MR-EGGER | ZHOU | 62 | 0.775 | 0.332 | 1.811 | -0.254 | 0.433 | 5.57e-01 | 46.48 | 0.900 | 0.031 | 2.66e-01 | - |
| MR-WMEDIAN | ZHOU | 62 | 1.112 | 0.868 | 1.425 | 0.106 | 0.126 | 4.01e-01 | - | - | - | - | - |
| MR-MBE | ZHOU | 62 | 0.960 | 0.521 | 1.770 | -0.040 | 0.312 | 8.97e-01 | - | - | - | - | - |
| MR-PRESSO | ZHOU | 62 | 1.242 | 1.064 | 1.450 | 0.217 | 0.079 | 7.77e-03 | - | - | - | - | 0 |
| IVW-MR | NEALE | 62 | 0.992 | 0.988 | 0.996 | -0.008 | 0.002 | 1.70e-05 | 102.659 | 0.001 | - | - | - |
| MR-EGGER | NEALE | 62 | 1.002 | 0.984 | 1.021 | 0.002 | 0.009 | 7.91e-01 | 100.343 | 0.001 | -0.001 | 2.39e-01 | - |
| MR-WMEDIAN | NEALE | 62 | 0.991 | 0.987 | 0.996 | -0.009 | 0.002 | 1.19e-04 | - | - | - | - | - |
| MR-MBE | NEALE | 62 | 0.993 | 0.981 | 1.005 | -0.007 | 0.006 | 2.40e-01 | - | - | - | - | - |
| MR-PRESSO | NEALE | 62 | 0.992 | 0.988 | 0.996 | -0.008 | 0.002 | 6.24e-05 | - | - | - | - | 0 |
| IVW-MR | MRCIEU | 62 | 1.010 | 1.006 | 1.014 | 0.010 | 0.002 | 1.74e-07 | 138.608 | 0 | - | - | - |
| MR-EGGER | MRCIEU | 62 | 0.997 | 0.979 | 1.015 | -0.003 | 0.009 | 7.45e-01 | 134.003 | 0 | 0.001 | 1.51e-01 | - |
| MR-WMEDIAN | MRCIEU | 62 | 1.012 | 1.008 | 1.016 | 0.012 | 0.002 | 1.25e-08 | - | - | - | - | - |
| MR-MBE | MRCIEU | 62 | 1.015 | 1.003 | 1.026 | 0.015 | 0.006 | 1.23e-02 | - | - | - | - | - |
| MR-PRESSO | MRCIEU | 61 | 1.011 | 1.007 | 1.014 | 0.011 | 0.002 | 3.77e-07 | - | - | - | - | 1 |
| IVW-MR | FinnGenR5 | 62 | 1.130 | 0.940 | 1.358 | 0.122 | 0.094 | 1.94e-01 | 85.041 | 0.023 | - | - | - |
| MR-EGGER | FinnGenR5 | 62 | 0.764 | 0.316 | 1.845 | -0.270 | 0.450 | 5.49e-01 | 83.933 | 0.022 | 0.026 | 3.73e-01 | - |
| MR-WMEDIAN | FinnGenR5 | 62 | 1.063 | 0.843 | 1.339 | 0.061 | 0.118 | 6.07e-01 | - | - | - | - | - |
| MR-MBE | FinnGenR5 | 62 | 1.000 | 0.559 | 1.788 | 0.000 | 0.297 | 9.99e-01 | - | - | - | - | - |
| MR-PRESSO | FinnGenR5 | 61 | 1.087 | 0.917 | 1.289 | 0.083 | 0.087 | 3.41e-01 | - | - | - | - | 1 |
| IVW-MR | FinnGenR9 | 62 | 1.073 | 0.958 | 1.202 | 0.070 | 0.058 | 2.24e-01 | 71.442 | 0.170 | - | - | - |
| MR-EGGER | FinnGenR9 | 62 | 1.254 | 0.728 | 2.161 | 0.226 | 0.278 | 4.15e-01 | 71.051 | 0.156 | -0.01 | 5.66e-01 | - |
| MR-WMEDIAN | FinnGenR9 | 62 | 1.128 | 0.968 | 1.315 | 0.120 | 0.078 | 1.24e-01 | - | - | - | - | - |
| MR-MBE | FinnGenR9 | 62 | 1.149 | 0.770 | 1.714 | 0.139 | 0.204 | 4.95e-01 | - | - | - | - | - |
| MR-PRESSO | FinnGenR9 | 62 | 1.073 | 0.958 | 1.202 | 0.070 | 0.058 | 2.29e-01 | - | - | - | - | 0 |
| IVW-MR | FinnGenR10 | 62 | 1.114 | 1.000 | 1.240 | 0.108 | 0.055 | 4.92e-02 | 74.027 | 0.122 | - | - | - |
| MR-EGGER | FinnGenR10 | 62 | 1.325 | 0.792 | 2.218 | 0.282 | 0.263 | 2.84e-01 | 73.468 | 0.114 | -0.011 | 4.99e-01 | - |
| MR-WMEDIAN | FinnGenR10 | 62 | 1.143 | 0.987 | 1.323 | 0.133 | 0.075 | 7.38e-02 | - | - | - | - | - |
| MR-MBE | FinnGenR10 | 62 | 1.244 | 0.833 | 1.859 | 0.219 | 0.205 | 2.86e-01 | - | - | - | - | - |
| MR-PRESSO | FinnGenR10 | 62 | 1.114 | 1.000 | 1.240 | 0.108 | 0.055 | 5.38e-02 | - | - | - | - | 0 |
| IVW-MR | SAIGE | 62 | 1.16 | 1.091 | 1.233 | 0.148 | 0.031 | 2.15e-06 | 99.075 | 0.001 | - | - | - |
| MR-EGGER | SAIGE | 62 | 1.089 | 0.807 | 1.47 | 0.085 | 0.153 | 5.78e-01 | 98.784 | 0.001 | 0.004 | 6.74e-01 | - |
| MR-WMEDIAN | SAIGE | 62 | 1.162 | 1.077 | 1.253 | 0.15 | 0.039 | 9.83e-05 | - | - | - | - | - |
| MR-MBE | SAIGE | 62 | 1.284 | 1.018 | 1.62 | 0.25 | 0.119 | 3.51e-02 | - | - | - | - | - |
| MR-PRESSO | SAIGE | 62 | 1.16 | 1.091 | 1.233 | 0.148 | 0.031 | 1.33e-05 | - | - | - | - | 0 |
| IVW-MR | PLINK | 62 | 1.175 | 1.098 | 1.257 | 0.161 | 0.035 | 3.11e-06 | 114.679 | 0 | - | - | - |
| MR-EGGER | PLINK | 62 | 1.139 | 0.817 | 1.588 | 0.13 | 0.169 | 4.42e-01 | 114.612 | 0 | 0.002 | 8.52e-01 | - |
| MR-WMEDIAN | PLINK | 62 | 1.194 | 1.103 | 1.293 | 0.177 | 0.041 | 1.22e-05 | - | - | - | - | - |
| MR-MBE | PLINK | 62 | 1.322 | 1.045 | 1.672 | 0.279 | 0.12 | 2.00e-02 | - | - | - | - | - |
| MR-PRESSO | PLINK | 61 | 1.163 | 1.089 | 1.241 | 0.151 | 0.033 | 3.07e-05 | - | - | - | - | 1 |

**Supplementary Table S3. Full MR results for analyses examining the effect of myopia on POAG.**

| Method | Sum. stats | Num. IVs | OR | LCI95 | UCI95 | BETA | SE | P | Qstat | Qpval | Egger intercept | Egger P | Num. MRP outliers |
|---|---|---|---|---|---|---|---|---|---|---|---|---|---|
| IVW-MR | JIANG | 32 | 1.142 | 1.040 | 1.253 | 0.132 | 0.048 | 5.56e-03 | 41.162 | 0.105 | - | - | - |
| MR-EGGER | JIANG | 32 | 1.482 | 1.104 | 1.988 | 0.393 | 0.150 | 8.78e-03 | 37.042 | 0.176 | -0.018 | 6.78e-02 | - |
| MR-WMEDIAN | JIANG | 32 | 1.128 | 0.998 | 1.275 | 0.120 | 0.063 | 5.46e-02 | - | - | - | - | - |
| MR-MBE | JIANG | 32 | 1.113 | 0.874 | 1.417 | 0.107 | 0.123 | 3.85e-01 | - | - | - | - | - |
| MR-PRESSO | JIANG | 32 | 1.142 | 1.040 | 1.253 | 0.132 | 0.048 | 9.33e-03 | - | - | - | - | 0 |
| IVW-MR | ZHOU | 1 | 1.005 | 0.899 | 1.125 | 0.005 | 0.057 | 9.26e-01 | - | - | - | - | - |
| MR-EGGER | - | - | - | - | - | - | - | - | - | - | - | - | - |
| MR-WMEDIAN | - | - | - | - | - | - | - | - | - | - | - | - | - |
| MR-MBE | - | - | - | - | - | - | - | - | - | - | - | - | - |
| IVW-MR | NEALE | 17 | 0.066 | 0.018 | 0.242 | -2.718 | 0.663 | 4.14e-05 | 16.096 | 0.446 | - | - | - |
| MR-EGGER | NEALE | 17 | 0.032 | 0.000 | 2.875 | -3.452 | 2.300 | 1.33e-01 | 15.977 | 0.384 | 0.004 | 7.38e-01 | - |
| MR-WMEDIAN | NEALE | 17 | 0.044 | 0.007 | 0.284 | -3.131 | 0.956 | 1.05e-03 | - | - | - | - | - |
| MR-MBE | NEALE | 17 | 0.203 | 0.007 | 6.175 | -1.595 | 1.743 | 3.60e-01 | - | - | - | - | - |
| MR-PRESSO | NEALE | 17 | 0.066 | 0.018 | 0.242 | -2.718 | 0.663 | 8.37e-04 | - | - | - | - | 0 |
| IVW-MR | MRCIEU | 36 | 8.977 | 2.188 | 36.829 | 2.195 | 0.720 | 2.31e-03 | 63.126 | 0.002 | - | - | - |
| MR-EGGER | MRCIEU | 36 | 206.8 | 2.566 | 16672.69 | 5.332 | 2.240 | 1.73e-02 | 59.321 | 0.005 | -0.015 | 1.40e-01 | - |
| MR-WMEDIAN | MRCIEU | 36 | 7.041 | 1.394 | 35.557 | 1.952 | 0.826 | 1.82e-02 | - | - | - | - | - |
| MR-MBE | MRCIEU | 36 | 10.13 | 0.577 | 177.963 | 2.316 | 1.462 | 1.13e-01 | - | - | - | - | - |
| MR-PRESSO | MRCIEU | 35 | 7.124 | 1.932 | 26.271 | 1.963 | 0.666 | 5.73e-03 | - | - | - | - | 1 |
| IVW-MR | FinnGenR9 | 5 | 1.108 | 0.992 | 1.238 | 0.102 | 0.057 | 7.04e-02 | 8.3 | 0.081 | - | - | - |
| MR-EGGER | FinnGenR9 | 5 | 0.833 | 0.671 | 1.035 | -0.183 | 0.111 | 9.88e-02 | 0.705 | 0.872 | 0.062 | 5.85e-03 | - |
| MR-WMEDIAN | FinnGenR9 | 5 | 1.149 | 1.031 | 1.281 | 0.139 | 0.055 | 1.20e-02 | - | - | - | - | - |
| MR-MBE | FinnGenR9 | 5 | 1.142 | 0.972 | 1.341 | 0.133 | 0.082 | 1.06e-01 | - | - | - | - | - |
| MR-PRESSO | FinnGenR9 | 5 | 1.108 | 0.992 | 1.238 | 0.102 | 0.057 | 1.45e-01 | - | - | - | - | 0 |
| IVW-MR | FinnGenR10 | 8 | 1.059 | 0.966 | 1.162 | 0.058 | 0.047 | 2.22e-01 | 11.148 | 0.132 | - | - | - |
| MR-EGGER | FinnGenR10 | 8 | 0.816 | 0.679 | 0.982 | -0.203 | 0.094 | 3.14e-02 | 2.093 | 0.911 | 0.059 | 2.62e-03 | - |
| MR-WMEDIAN | FinnGenR10 | 8 | 1.033 | 0.933 | 1.144 | 0.033 | 0.052 | 5.29e-01 | - | - | - | - | - |
| MR-MBE | FinnGenR10 | 8 | 1.022 | 0.892 | 1.171 | 0.022 | 0.069 | 7.53e-01 | - | - | - | - | - |
| MR-PRESSO | FinnGenR10 | 8 | 1.059 | 0.966 | 1.162 | 0.058 | 0.047 | 2.62e-01 | - | - | - | - | 0 |
| IVW-MR | SAIGE | 48 | 1.119 | 1.033 | 1.213 | 0.113 | 0.041 | 5.72e-03 | 147.671 | 0 | - | - | - |
| MR-EGGER | SAIGE | 48 | 1.221 | 0.996 | 1.496 | 0.199 | 0.104 | 5.48e-02 | 145.071 | 0 | -0.009 | 3.64e-01 | - |
| MR-WMEDIAN | SAIGE | 48 | 1.119 | 1.039 | 1.205 | 0.113 | 0.038 | 2.88e-03 | - | - | - | - | - |
| MR-MBE | SAIGE | 48 | 1.105 | 0.985 | 1.239 | 0.099 | 0.059 | 9.01e-02 | - | - | - | - | - |
| MR-PRESSO | SAIGE | 45 | 1.126 | 1.064 | 1.192 | 0.119 | 0.029 | 1.82e-04 | - | - | - | - | 3 |
| IVW-MR | PLINK | 54 | 1.105 | 1.028 | 1.188 | 0.100 | 0.037 | 6.47e-03 | 132.248 | 0 | - | - | - |
| MR-EGGER | PLINK | 54 | 1.208 | 0.980 | 1.490 | 0.189 | 0.107 | 7.68e-02 | 130.268 | 0 | -0.009 | 3.74e-01 | - |
| MR-WMEDIAN | PLINK | 54 | 1.106 | 1.027 | 1.192 | 0.101 | 0.038 | 8.07e-03 | - | - | - | - | - |
| MR-MBE | PLINK | 54 | 1.109 | 0.975 | 1.262 | 0.103 | 0.066 | 1.16e-01 | - | - | - | - | - |
| MR-PRESSO | PLINK | 50 | 1.113 | 1.054 | 1.175 | 0.107 | 0.028 | 3.40e-04 | - | - | - | - | 4 |

**Figure S1. Graphs of SNP vs. EduYears and SNP vs. Myopia regression coefficients for the inverse variance-weighted Mendelian randomization analyses using different sets of myopia summary statistics.** The blue line is the inverse-variance weighted random effects model fit. Error bars indicate 95% confidence intervals.

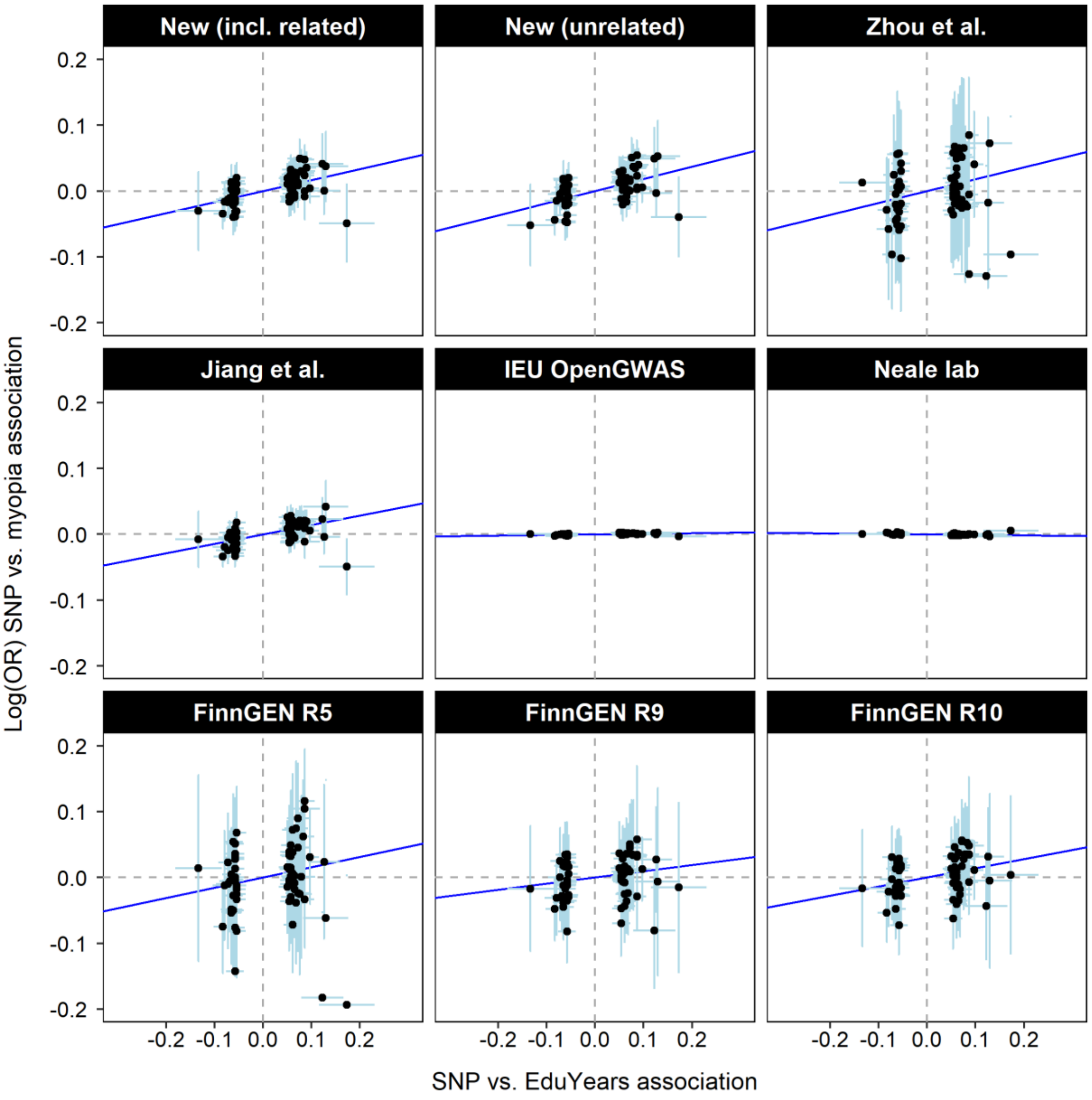

**Figure S2. Graphs of SNP vs. Myopia and SNP vs. POAG regression coefficients for the inverse variance-weighted Mendelian randomization analyses using different sets of myopia summary statistics.** The blue line is the inverse-variance weighted random effects model fit. Error bars indicate 95% confidence intervals. The number of SNPs (instrumental variables) varies for each set of summary statistics, depending on the number of independent, genome-wide significant SNPs.

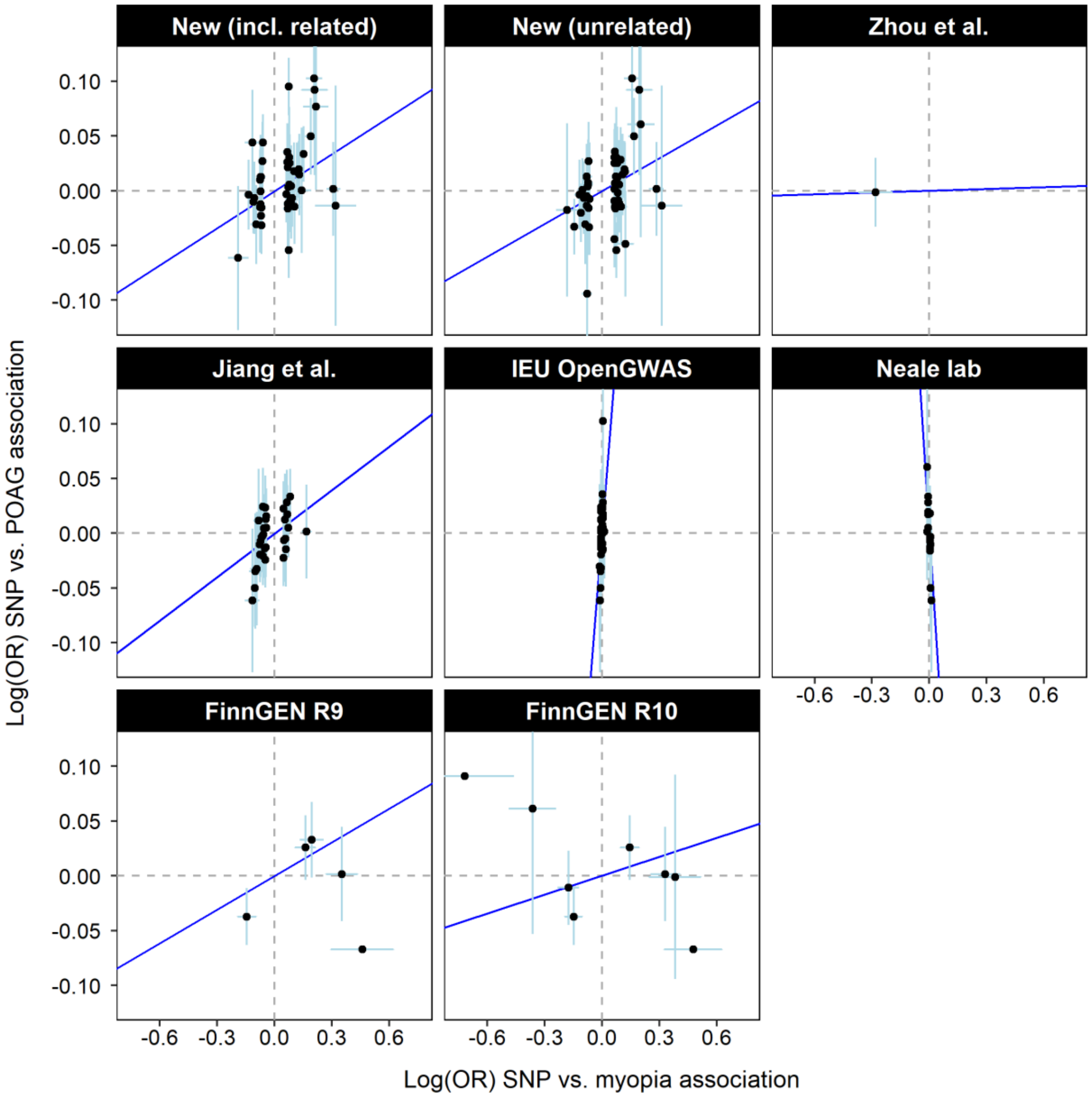
